# A hospital-based cluster-randomized controlled trial of free influenza vaccination before hospital discharge compared to routine care in patients with heart failure (PANDA II): Statistical Analysis Plan

**DOI:** 10.1101/2024.12.20.24319430

**Authors:** Sana Shan, Laurent Billot, Xin Du, Jianzeng Dong, Craig Anderson

**Affiliations:** The George Institute for Global Health, UNSW Sydney, Australia; Heart Health Research Center, Anzhen Hospital, Beijing, China; Beijing Anzhen Hospital, Capital Medical University, Beijing, China

**Keywords:** Cardiovascular Medicine, Heart Failure, Influenza, Vaccination, Clinical Trial, Cluster randomized trial, Statistical Analysis Plan

## Abstract

The PANDA II trial aims to evaluate whether a free influenza vaccination program for patients hospitalized in China with acute heart failure provides benefits compared to current routine practice of no influenza vaccination. It is designed as a two-arm, parallel, cluster-crossover randomized trial of 122 hospitals conducted over three influenza seasons. This statistical analysis plan pre-specifies the method of analysis for every outcome and key variable collected in the trial.

The primary outcome is a composite of all-cause mortality and re-admission to hospital within a 12- month period after the initial 30-day post-discharge period. The primary analysis will include all recruited participants analysed according to the intervention allocated to the hospital during their enrolment period. It will consist in an individual-level hierarchical logistic regression adjusted for the effect of the period and for clustering by hospital using both a random hospital effect and a random hospital-period effect. The effect of the influenza vaccination program will be estimated as an odds ratio and 95% confidence interval.

Sensitivity analyses will include covariate adjustments as well as a cluster-level analysis and subgroup analyses according to eight pre-specified subgroups.

## 1 Administrative information

### 1.1 Study identifiers

Chinese Clinical Trial Registry, ChiCTR.org.cn register Identifier: ChiCTR2100053264, Date: 17 November 2021

### 1.2 Revision history

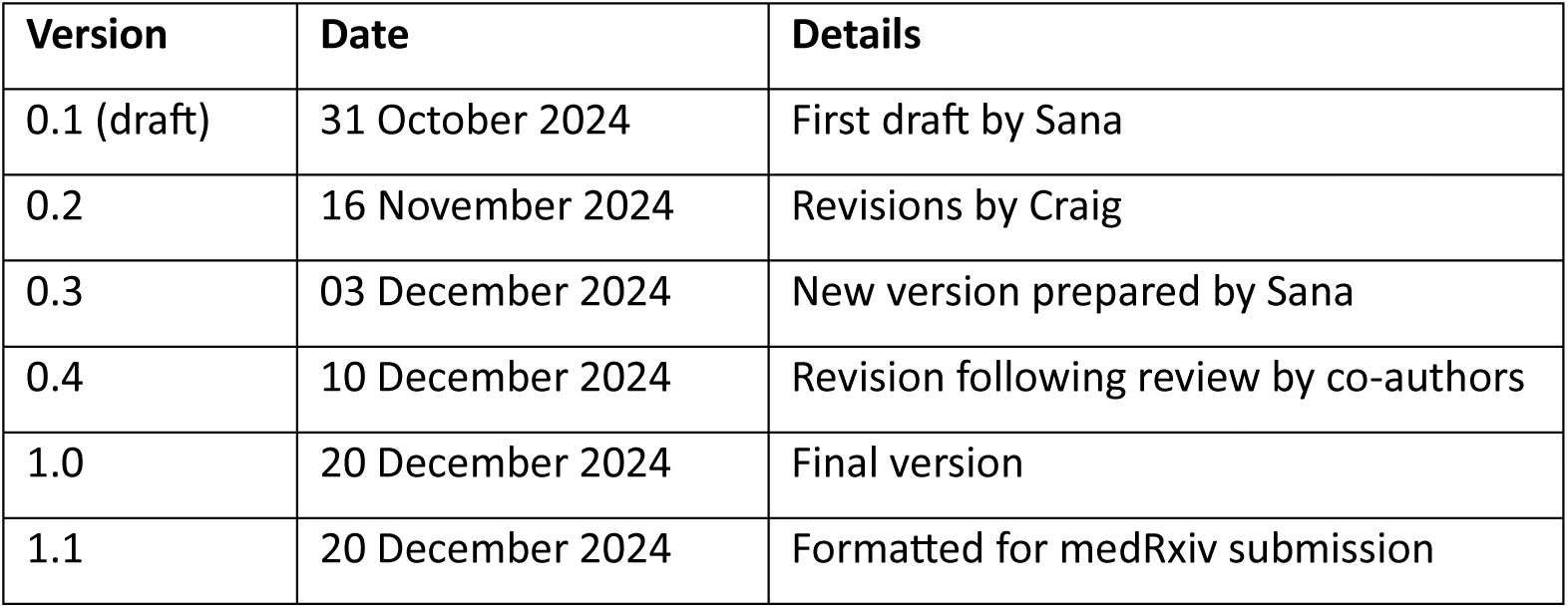

| Version | Date | Details |
| --- | --- | --- |
| 0.1 (draft) | 31 October 2024 | First draft by Sana |
| 0.2 | 16 November 2024 | Revisions by Craig |
| 0.3 | 03 December 2024 | New version prepared by Sana |
| 0.4 | 10 December 2024 | Revision following review by co-authors |
| 1.0 | 20 December 2024 | Final version |
| 1.1 | 20 December 2024 | Formatted for medRxiv submission |

### 1.3 Contributors to the statistical analysis plan

SS – Main author; developed the initial draft and prepared subsequent versions. LB, CA - Reviewed every draft and revisions. XD, JD - Reviewed and approved final draft.

## 2 Introduction

### 2.1 Study synopsis

The PANDA II trial ^(1)^ is a two-arm, parallel, cluster-randomised controlled trial designed to evaluate whether free influenza vaccination program for patients hospitalized with acute heart failure (HF) provides benefits compared to current routine practice of no influenza vaccination. In this trial, the unit of the cluster is the hospital. There are three one-year study periods that correspond to the winter influenza seasons, with the potential for participating hospitals to be re-randomised each year. In the first year, due to the impact of COVID-19, the patient enrolment schedule was determined according to influenza activity in the community: enrolment started when the population influenza infection rate reached 3%, according to the monitoring report prepared by the Centers for Disease Control and Prevention (CDC) in China, and enrolment took place between December 3, 2021 and January 30, 2022. In the second year, it occurred between September 1, 2022 and January 20, 2023. In the third year, the enrolment period was from September 4, 2023 to February 8, 2024. Enrolment was to stop when the target quota of 50 patients per study site was reached. Patients have been followed up for 12-months after their separation/discharge from hospital. Study hospitals enrolled consecutive HF patients during each recruitment window. Hospitals allocated to the intervention group have had a point of vaccination (POV) established on the HF wards to provide free influenza vaccination to patients immediately prior to their discharge from hospital. Hospitals allocated to the control group do not provide routine influenza vaccination but have this recommended to patients upon hospital discharge through a community health care centre.

### 2.2 Study population

There are 93 county-level, mainly secondary, hospitals that are geographically dispersed across China have been included in the study. These hospitals are located in the provinces of Henan, Jilin, Hebei, Shaanxi, Hunan, and Guangxi. They have been randomly assigned to either the intervention or control arm in each year.^(1)^ A total of 169 clusters have been randomized over the 3 years of the study.

### 2.2.1 Inclusion Criteria

#### Hospital level

Hospital sites are eligible if they meet all following requirement:

- able to admit and treat HF patients
- capable to enrol 50 continuous HF patients during each recruitment window period
- have staff available to work as part of a study team in research
- have local administration approval to provide influenza vaccination in the hospital ward

#### Individual level

Patients who present to participating centers are eligible if they meet all the following requirement:

- adults (age ≥18 years)
- have been admitted to hospital during the study recruitment period
- Have a diagnosis of HF with a New York Heart Association (NYHA) classification of functional severity of III (marked limitation in activity due to symptoms, even during less-than-ordinary activity, e.g. walking short distances 20-100 meters) or IV (severe limitations - experiences symptoms even while at rest)
- Consecutively recruited within the quota of 50 patients for each site
- Written consent to participate in the study

### 2.2.2 Exclusion Criteria

#### Hospital level

Hospital sites are not eligible if they meet any of the following criteria:

- Participating in any other influenza vaccination-related programs that might confound this study
- Free influenza vaccination policy has already been implemented for local residents

#### Individual level

Patients are not eligible if they meet any of the following criteria:

- previous enrolment in this study
- hospitalised for HF ≥2 times in the past two months
- known allergy to influenza vaccine
- had a COVID vaccination in the past 14 days
- discharge from hospital with the intention of being transferred to another medical facility for further treatment.

## 3 Study interventions

Hospitals allocated to the intervention arm will establish a point of vaccination (POV) in the cardiovascular wards to provide free influenza vaccination to patients. As part of the intervention, healthcare teams in the intervention group will be educated in the process of vaccination to enhance their willingness to vaccinate patients with HF. Patients eligible for this study in the intervention group who consent to follow up will be enrolled regardless of their wiliness to be vaccinated. Routine standard of care practice was to be maintained in both groups of hospitals, but no vaccination intervention was to occur in the control group hospitals in the study recruitment period. In the control arm hospitals, doctors can suggest to patients that they can receive influenza vaccination at community health care centre according to usual care.

### 3.1 Randomisation

The randomisation unit for intervention or control groups is the hospital. Randomisation was performed in each of three influenza seasons. Computer-generated random numbers were used to assign participating hospitals to the intervention or control group in a 1:1 ratio, stratified by province to ensure the numbers of intervention and control hospitals were balanced. Site investigators remained blind to the treatment allocation until they were notified on the study group in which they were assigned at about one month before any patients were enrolled.

### 3.2 Study treatment

All consenting patients in the intervention group were given the Sanofi Pasteur trivalent (<10%) or other Chinese manufactured quadrivalent (∼90%) inactivated influenza vaccine (0.5 ml) prior to their discharge from hospital.

## 4 Outcomes

### 4.1 Primary outcome

The primary outcome will comprise the following events that occur in the 12 months period after the initial 30 days post-discharge period, and excluding events occurring during the non-influenza circulation season (defined as from June 1 to August 31 in northern China):

- the composite of all-cause mortality and re-admission to hospital

The reason for censoring events in the first 30 days after discharge from hospital is that these are presumed to be more likely related to cardiac disease that is less likely to be prevented from influenza vaccination. In north China (Heilongjiang, Liaoning, Jilin, Hebei, Shaanxi, Shanxi, Shandong) the period from June 1 to August 31 is considered the non-influenza season, during which events are unlikely to be attributed to influenza infection. In South China (Henan, Hunan, and Guangxi), the general experience is for two peak influenza seasons each year, one in summer and the other in winter. Thus, we did not define a non-influenza season for the Southern provinces.

### 4.2 Secondary outcomes

The secondary outcomes are the following events (excluding the 30-day post-discharge period)::

- all-cause mortality within 12 months
- all-cause readmission to hospital within 12 months
- all-cause mortality within 6 months
- all-cause readmission within 6 months
- composite of all-cause mortality or readmission within 6 months

### 4.3 Exploratory outcomes

- number of hospitals readmissions over 12 months (hospitalisations for any cause)
- number of infectious-like illnesses (ILI), defined as ≥38 degrees Celsius, cough or sore throat, in the previous 7 days at each follow-up assessment

### 4.4 Safety outcomes

- any serious adverse event (SAE), all-cause and cause-specific

## 5 Analysis principles

### 5.1 Sample size

A total of 122 hospitals participating over 3 influenza seasons in which 50 patients are recruited in each hospital will provide 90% power and a two-tailed significance level of <0.05 to detect a significant 13% reduction effect for the primary outcome in the intervention group on the assumption that the 12-month mortality or hospital readmissions rate for patients with HF in the control group is expected to be 50 per 100 person-years, after excluding events in non-influenza season or within 30 days after discharge. Other assumptions are: that at least 90% of the intervention group will receive the influenza vaccine; that 10% of those in the control group will receive influenza vaccine; 15% of the population will develop influenza in the winter; influenza vaccination will protect 60% of individuals from influenza infection; influenza infection in severe HF patients will lead 80% of this population to experience a re-hospitalization or death; an intra-class correlation coefficient (ICC) of 0.03 across participating hospitals; and a 12 month follow-up dropout of 10%.

### 5.2 Software

Analyses will be conducted primarily using SAS Enterprise Guide (version 8.3 or above) and R (version 4.0.0 or above).

### 5.3 Interim analyses

No formal interim analyses were conducted during the study.

### 5.4 Multiplicity adjustment

Statistical tests are to be two-sided with a nominal level of *α* set at 5%. Analysis of the primary composite outcome will be unadjusted for multiplicity. For the 5 secondary outcomes measured, we will control the family-wise error rate by applying a sequential Holm-Sidak correction ^(2)^. All p-values will be ranked from smallest to largest, and then comparing them to an adjusted level of significance calculated as 1-(1-0.05),1/C where C indicates the number of comparisons that remain. The sequential testing procedure stops as soon as a p value fails to reach the corrected significance level.

### 5.5 Change in the statistical analysis plan from the protocol

As the study progressed over three years with difference in virulence including Covid, a decision was made to change the analysis of the primary outcome from a cluster-level analysis to an individual-level analysis to adjust for correlations between patients within the same cluster, same cluster and different time periods as well as adjust for individual baseline characteristics.

Cluster-level analysis of the primary outcome has now been added as a sensitivity analysis.

### 5.6 Datasets analysed

#### 5.6.1 Analysis populations

Due to the cluster design of the study with re-randomisation of cluster hospitals at each of the three intervention periods, the group allocation for a patient is determined by the site and by the period during their participation, regardless of treatment adherence. The intention-to-treat (ITT) analysis set will be used to assess both effectiveness and safety.

#### 5.6.2 Analysis strategy

For all outcomes, the analyses will be performed on the ITT population using all available data. It is expected that the rate of missingness will be less than 3%, therefore, no imputation is planned for missing data.

For participants lost to follow-up, all available information until the time of death or loss to follow-up will be used in analysis.

## 6 Planned Analyses

### 6.1 Subject disposition

The flow of patients through the trial will be displayed in a CONSORT^(3)^ (Consolidated Standards of Reporting Trials) diagram [see Figure 1]. The report will include the following: the number of centers assessed for eligibility, reasons for exclusion, the number of enrolled participants, and the proportion of eligible patients being enrolled in each treatment group of the study for each of the three study periods. The flowchart will also include the number of patients alive and available at end of follow-up (12-Month) for each of the three study periods (1^st^ year, 2^nd^ year and 3^rd^ year) as well as the number of patients lost to follow-up.

### 6.2 Baseline comparison

#### 6.2.1 Cluster characteristics

Description of the cluster characteristics (e.g. province, no. of beds (above and below median values), annual number of patients admitted with HF (above and below median values) will be presented by treatment group. Discrete variables will be summarised by frequencies and percentages. Percentages will be calculated according to the number of clusters with available data. Continuous variables will be summarised by using mean and standard deviation (SD), and median and interquartile range (Q1-Q3).

#### 6.2.2 Patient characteristics

Description of baseline patient characteristics will be presented by treatment group. Discrete variables will be summarised by frequencies and percentages. Percentages will be calculated according to the number of patients in whom data are available. Continuous variables will be summarised by using mean and SD, and median and interquartile range (Q1-Q3). No adjustment for clustering will be applied when summarising baseline characteristics. Baseline measures for all patients will be tabulated for the variables listed below

- Anthropometrics: age (years), sex, weight (kg), height (cm) and BMI (kg/m^2^)
- Demographics and socio-economic status including ethnicity, education, marital status, occupation, type of insurance (hospital expenses)
- Combined prior medical conditions (from admission-baseline registration form)
- Diagnostic methods incl. echocardiography, electrocardiogram, chest radiograph, coronary angiography
- Lifestyle factors; smoking and alcohol (from admission-baseline registration form)
- New York Heart Association NYHA functional class (baseline registration form)
- Concomitant medication; antibiotics, IV inotropic drugs, IV vasodilators, infusion therapy (from admission registration form)
- Interventional in-hospital treatments including permanent pacemaker insertion, implantable cardioverter defibrillator (ICD), cardiac resynchronization (CRT), percutaneous coronary intervention (PCI), other percutaneous procedures
- EQ-5D assessment at the time of admission
- Clinical data (first laboratory examination results after admission)
- Vital signs (systolic BP, diastolic BP, heart rate)

### 6.3 Follow-up assessments

All assessments performed and interventions received during follow-up will be described by treatment group. Details are presented in mock tables. No formal statistical tests are planned for these variables.

### 6.4 Analysis of the primary outcome

#### Main analysis (Individual-level)

Individual-level analysis will be performed to summarise the composite of all-cause mortality and all-cause hospitalisation in the 12-month post-discharge follow-up period, excluding events which occurred within 30 days of discharge or in the non-influenza season period for the northern hospitals (as described in detail in section 5.1). A binary variable indicating whether each patient experienced the composite primary endpoint within the given timeframe will be created. The primary analysis will consider two-levels of clustering: 1) clustering of participants from the same hospital and same period (Year 1, 2 or 3) and 2) clustering of participants from the same hospital but different period. The intervention effect will be estimated using a hierarchical logistic regression model with a binomial distribution and logit link function. The model will include a fixed effect of intervention (influenza vaccine vs control), a random cluster effect for hospital and a random hospital-period effect. The results will be presented as odds ratio (OR) with 95% confidence intervals (CI).

### 6.5 Adjusted analyses

Adjusted analyses of the primary outcome will be performed using individual patient-level data by adding the following baseline covariates to the main analysis model (see section 7.4): age, NTHC, sex, NYHC severity, history of previous history of ischaemic heart disease [history of myocardial infarction, coronary stent insertion, CAVG, or coronary artery disease; Yes vs. No], LVEF (<40% vs ≥40%), renal function (eGFR <30 vs ≥30 mL/mL/min/1.73m^2^. The adjusted treatment effect will be reported as the odds ratio (OR) and 95% CI.

### 6.6 Sensitivity analyses

As part of the sensitivity analysis, a cluster-level analysis will be performed to summarise the primary outcome of composite rate of all-cause death and all-cause hospitalization in influenza season within first year of discharge (excluding events which are within 30 days of discharge or in the non-influenza season). The proportion of participants experiencing a primary outcome will be calculated for each hospital and for each of the 3 periods. Summary measures in each cluster-period will then be modelled using the following linear model (see equation 13 in Morgan et al^(4)^)

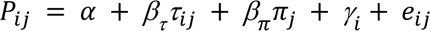

where *P*_*ij*_ is the proportion of events in period j of cluster 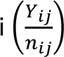, *α* is the overall intercept, *β_τ_* is the treatment effect corresponding to the average difference in proportions and *τ*_*ij*_ is an indicator variable for treatment of interest, *β*_*π*_ is a fixed period effect and *π*_*j*_ is an indicator variable for period, *γ*_*i*_ is a fixed effect of cluster, *e*_*ij*_ is a normally distributed residual error term with mean zero and variance *σ*_2*e*_

The regression will be weighted proportionally to the inverse of the binomial variance for each cluster-period with each weight calculated as:

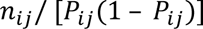

### 6.7 Treatment of missing data

We are not planning to conduct any multiple imputation for primary outcome. See Section 7.4 for more details about the main analysis.

### 6.8 Per-protocol analysis

No per-protocol analysis will be performed.

### 6.9 Analysis of the secondary outcomes

#### Individual-level analysis

Individual-level analyses will be performed for all 5 secondary outcomes. Given the binary nature of the outcomes, the same analysis approach will be applied as the one used for the primary outcome main analysis, see section 7.4 for details. No adjusted or cluster-level analysis is planned for secondary outcomes.

### 6.10 Analysis of the exploratory outcomes

Analysis of exploratory outcomes including number of hospital readmissions for any cause over one year period, and incidence of ILI will be performed using linear regression with exploratory outcome collected over 12-month follow-up period. The model will include a fixed effect of the intervention (influenza vaccine, control), a random cluster effect for hospital and a random hospital-period effect. The effect of the intervention will be presented as mean difference and its 95% CI, using the control arm as the reference.

### 6.11 Analysis of safety outcomes

Safety outcomes of all-cause and cause-specific SAEs will be summarised as the number and proportion of patients experiencing at least one event. Any clinical sign or symptom other than the defined endpoint that are unexpected, will be reported within 7 days post-vaccination. Adverse events will be summarised by category of event and overall numbers of events. In addition to the number of patients with at least one event, we will report the total number of events. Comparison of total number of adverse events between treatment arms will involve obtaining cluster level mean rates using linear regression summary measures (see section 7.6 Sensitivity analysis for further details)

A listing of all AEs and SAEs will be reported (in an appendix).

### 6.12 Subgroup analyses

Heterogeneity of treatment on the primary endpoint will be assessed in the following pre-defined subgroups:

- age category (<70 vs. ≥70 years)
- sex
- province of hospital location (North vs. Central vs. South)
- year of treatment (1^st^ year, 2^nd^ year, 3^rd^ year)
- severity of HF (NYHA classification)
- history of coronary artery disease
- LVEF (<40% vs. ≥40%)
- level of renal function (eGFR <30 vs ≥30 mL/mL/min/1.73m2)

The analysis for each subgroup will be performed by adding the subgroup variable as well as its interaction with the intervention as fixed effects to the main analysis individual-level model for primary outcome, as described in section 7.4. Within each subgroup, summary measures will include frequency of events and percentage, within each treatment arm, as well as the odds ratio (OR) for treatment effect with a 95% CI. The results will be displayed on a forest plot [figure 2] including the p-value for heterogeneity corresponding to the interaction term between the intervention and the subgroup variable.

## Data Availability

The trial is ongoing and no data is available yet.

## 8 Appendix 1: Proposed Tables and figures

**Figure 1:**
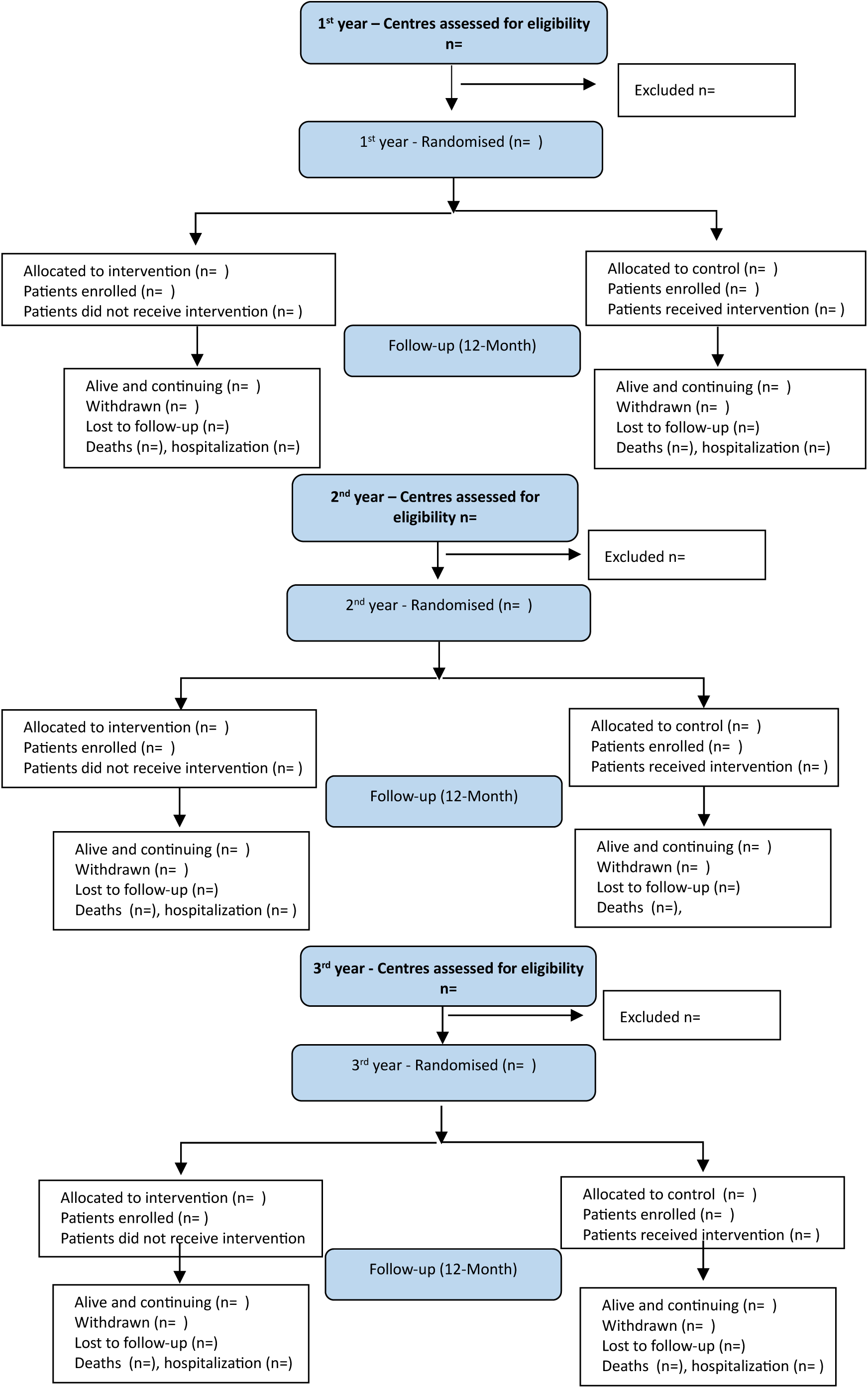
Consort flowchart.

**Table 1.** Enrolment by province, hospital and treatment period.

| Province/<br>Hospital | Year 1 |  | Year 2 |  | Year 3 |  | Total |  |
| --- | --- | --- | --- | --- | --- | --- | --- | --- |
|  | Screened | Enrolled | Screened | Enrolled | Screened | Enrolled | Screened | Enrolled |
| **Henan** | xx (xx.x%) | xx (xx.x%) | xx (xx.x%) | xx (xx.x%) | xx (xx.x%) | xx (xx.x%) | xx (xx.x%) | xx<br>(xx.x%) |
| Hospital 1 | xx (xx.x%) | xx (xx.x%) | xx (xx.x%) | xx (xx.x%) | xx (xx.x%) | xx (xx.x%) | xx (xx.x%) | xx<br>(xx.x%) |
| Hospital 2 | xx (xx.x%) | xx (xx.x%) | xx (xx.x%) | xx (xx.x%) | xx (xx.x%) | xx (xx.x%) | xx (xx.x%) | xx<br>(xx.x%) |
| etc... | xx (xx.x%) | xx (xx.x%) | xx (xx.x%) | xx (xx.x%) | xx (xx.x%) | xx (xx.x%) | xx (xx.x%) | xx<br>(xx.x%) |
| **Jilin** | xx (xx.x%) | xx (xx.x%) | xx (xx.x%) | xx (xx.x%) | xx (xx.x%) | xx (xx.x%) | xx (xx.x%) | xx<br>(xx.x%) |
| Hospital 1 | xx (xx.x%) | xx (xx.x%) | xx (xx.x%) | xx (xx.x%) | xx (xx.x%) | xx (xx.x%) | xx (xx.x%) | xx<br>(xx.x%) |
| Hospital 2 | xx (xx.x%) | xx (xx.x%) | xx (xx.x%) | xx (xx.x%) | xx (xx.x%) | xx (xx.x%) | xx (xx.x%) | xx<br>(xx.x%) |
| etc... | xx (xx.x%) | xx (xx.x%) | xx (xx.x%) | xx (xx.x%) | xx (xx.x%) | xx (xx.x%) | xx (xx.x%) | xx<br>(xx.x%) |
| **Hebei** | xx (xx.x%) | xx (xx.x%) | xx (xx.x%) | xx (xx.x%) | xx (xx.x%) | xx (xx.x%) | xx (xx.x%) | xx<br>(xx.x%) |
| Hospital 1 | xx (xx.x%) | xx (xx.x%) | xx (xx.x%) | xx (xx.x%) | xx (xx.x%) | xx (xx.x%) | xx (xx.x%) | xx<br>(xx.x%) |
| Hospital 2 | xx (xx.x%) | xx (xx.x%) | xx (xx.x%) | xx (xx.x%) | xx (xx.x%) | xx (xx.x%) | xx (xx.x%) | xx<br>(xx.x%) |
| etc... | xx (xx.x%) | xx (xx.x%) | xx (xx.x%) | xx (xx.x%) | xx (xx.x%) | xx (xx.x%) | xx (xx.x%) | xx<br>(xx.x%) |
| **Shaanxi** | xx (xx.x%) | xx (xx.x%) | xx (xx.x%) | xx (xx.x%) | xx (xx.x%) | xx (xx.x%) | xx (xx.x%) | xx<br>(xx.x%) |
| Hospital 1 | xx (xx.x%) | xx (xx.x%) | xx (xx.x%) | xx (xx.x%) | xx (xx.x%) | xx (xx.x%) | xx (xx.x%) | xx<br>(xx.x%) |
| Hospital 2 | xx (xx.x%) | xx (xx.x%) | xx (xx.x%) | xx (xx.x%) | xx (xx.x%) | xx (xx.x%) | xx (xx.x%) | xx<br>(xx.x%) |
| etc... | xx (xx.x%) | xx (xx.x%) | xx (xx.x%) | xx (xx.x%) | xx (xx.x%) | xx (xx.x%) | xx (xx.x%) | xx<br>(xx.x%) |
| **Hunan** | xx (xx.x%) | xx (xx.x%) | xx (xx.x%) | xx (xx.x%) | xx (xx.x%) | xx (xx.x%) | xx (xx.x%) | xx<br>(xx.x%) |
| Hospital 1 | xx (xx.x%) | xx (xx.x%) | xx (xx.x%) | xx (xx.x%) | xx (xx.x%) | xx (xx.x%) | xx (xx.x%) | xx<br>(xx.x%) |
| Hospital 2 | xx (xx.x%) | xx (xx.x%) | xx (xx.x%) | xx (xx.x%) | xx (xx.x%) | xx (xx.x%) | xx (xx.x%) | xx<br>(xx.x%) |
| etc... | xx (xx.x%) | xx (xx.x%) | xx (xx.x%) | xx (xx.x%) | xx (xx.x%) | xx (xx.x%) | xx (xx.x%) | xx (xx.x%) |
| Guangxi | xx (xx.x%) | xx (xx.x%) | xx (xx.x%) | xx (xx.x%) | xx (xx.x%) | xx (xx.x%) | xx (xx.x%) | xx (xx.x%) |
| Hospital 1 | xx (xx.x%) | xx (xx.x%) | xx (xx.x%) | xx (xx.x%) | xx (xx.x%) | xx (xx.x%) | xx (xx.x%) | xx (xx.x%) |
| Hospital 2 | xx (xx.x%) | xx (xx.x%) | xx (xx.x%) | xx (xx.x%) | xx (xx.x%) | xx (xx.x%) | xx (xx.x%) | xx (xx.x%) |
| etc... | xx (xx.x%) | xx (xx.x%) | xx (xx.x%) | xx (xx.x%) | xx (xx.x%) | xx (xx.x%) | xx (xx.x%) | xx (xx.x%) |

**Table 2:** Cluster characteristics.

| Hospital characteristics | Intervention | Control | Total |
| --- | --- | --- | --- |
| **Provincial location** | N=xxx | N=xxx | N=xxx |
| Xxx province | xx (xx.x%) | xx (xx.x%) | xx (xx.x%) |
| Xxx province | xx (xx.x%) | xx (xx.x%) | xx (xx.x%) |
| **Etc.** | xx (xx.x%) | xx (xx.x%) | xx (xx.x%) |
| **No. hospital beds** |  |  |  |
| N Mean(SD) | N xx.x (xx.x) | N xx.x (xx.x) | N xx.x (xx.x) |
| Median [Q1 – Q3] | xx.x [xx.x- xx.x] | xx.x [xx.x- xx.x] | xx.x [xx.x- xx.x] |
| **No. of beds above median** | xx (xx.x%) | xx (xx.x%) | xx (xx.x%) |
| **No. of beds below median** | xx (xx.x%) | xx (xx.x%) | xx (xx.x%) |
| **Annual number of patients admitted with HF** |  |  |  |
| N Mean(SD) | N xx.x (xx.x) | N xx.x (xx.x) | N xx.x (xx.x) |
| Median [Q1 – Q3] | xx.x [xx.x- xx.x] | xx.x [xx.x- xx.x] | xx.x [xx.x- xx.x] |
| **No. of patients admitted with HF (above median)** | xx (xx.x%) | xx (xx.x%) | xx (xx.x%) |
| **No. of patients admitted with HF (below median)** | xx (xx.x%) | xx (xx.x%) | xx (xx.x%) |

**Table 3:** Baseline characteristics table.

| Characteristics | Intervention | Control | Total |
| --- | --- | --- | --- |
| **Age (years)** |  |  |  |
| N Mean (SD) | N xx.x (xx.x) | N xx.x (xx.x) | N xx.x (xx.x) |
| Median [Q1 – Q3] | xx.x [xx.x- xx.x] | xx.x [xx.x- xx.x] | xx.x [xx.x- xx.x] |
| **Sex** | N=xxx | N=xxx | N=xxx |
| Male | xx (xx.x%) | xx (xx.x%) | xx (xx.x%) |
| Female | xx (xx.x%) | xx (xx.x%) | xx (xx.x%) |
| **Weight (kg)** |  |  |  |
| N Mean(SD) | N xx.x (xx.x) | N xx.x (xx.x) | N xx.x (xx.x) |
| Median [Q1 – Q3] | xx.x [xx.x- xx.x] | xx.x [xx.x- xx.x] | xx.x [xx.x- xx.x] |
| **BMI (kg/m<sup>2</sup>)** |  |  |  |
| N Mean(SD) | N xx.x (xx.x) | N xx.x (xx.x) | N xx.x (xx.x) |
| Median [Q1 – Q3] | xx.x [xx.x- xx.x] | xx.x [xx.x- xx.x] | xx.x [xx.x- xx.x] |
| **Ethnicity** | N=xxx | N=xxx | N=xxx |
| The Han nationality | xx (xx.x%) | xx (xx.x%) | xx (xx.x%) |
| The Hui nationality | xx (xx.x%) | xx (xx.x%) | xx (xx.x%) |
| The Man nationality | xx (xx.x%) | xx (xx.x%) | xx (xx.x%) |
| Other nationality | xx (xx.x%) | xx (xx.x%) | xx (xx.x%) |
| **Education** | N=xxx | N=xxx | N=xxx |
| Primary school and below | xx (xx.x%) | xx (xx.x%) | xx (xx.x%) |
| Secondary school | xx (xx.x%) | xx (xx.x%) | xx (xx.x%) |
| University and above | xx (xx.x%) | xx (xx.x%) | xx (xx.x%) |
| **Occupation** | N=xxx | N=xxx | N=xxx |
| Professionals (doctors, teachers, lawyers, etc.) | xx (xx.x%) | xx (xx.x%) | xx (xx.x%) |
| State civil servants or staff of public institutions | xx (xx.x%) | xx (xx.x%) | xx (xx.x%) |
| Sales and service personnel | xx (xx.x%) | xx (xx.x%) | xx (xx.x%) |
| Production personnel in agriculture/forestry/animal husbandry/fishery/water conservancy industry | xx (xx.x%) | xx (xx.x%) | xx (xx.x%) |
| Production, transportation, equipment operation personnel and related personnel | xx (xx.x%) | xx (xx.x%) | xx (xx.x%) |
| Military personnel | xx (xx.x%) | xx (xx.x%) | xx (xx.x%) |
| Private owners | xx (xx.x%) | xx (xx.x%) | xx (xx.x%) |
| Other persons inconvenient to classify | xx (xx.x%) | xx (xx.x%) | xx (xx.x%) |
| **Marital status** | N=xxx | N=xxx | N=xxx |
| Married | xx (xx.x%) | xx (xx.x%) | xx (xx.x%) |
| Divorced | xx (xx.x%) | xx (xx.x%) | xx (xx.x%) |
| Separated | xx (xx.x%) | xx (xx.x%) | xx (xx.x%) |
| Widowed | xx (xx.x%) | xx (xx.x%) | xx (xx.x%) |
| **Type of expenses** | N=xxx | N=xxx | N=xxx |
| Free medical treatment | xx (xx.x%) | xx (xx.x%) | xx (xx.x%) |
| Medical insurance for urban employees | xx (xx.x%) | xx (xx.x%) | xx (xx.x%) |
| Medical insurance for urban residents | xx (xx.x%) | xx (xx.x%) | xx (xx.x%) |
| Rural cooperative medical care | xx (xx.x%) | xx (xx.x%) | xx (xx.x%) |
| Self-paying | xx (xx.x%) | xx (xx.x%) | xx (xx.x%) |
| **Bad lifestyle habits** | N=xxx | N=xxx | N=xxx |
| History of heavy drinking in the past <sup>(1)</sup> | xx (xx.x%) | xx (xx.x%) | xx (xx.x%) |
| **Smoking status<sup>(2)</sup>** | N=xxx | N=xxx | N=xxx |
| Current smoker | xx (xx.x%) | xx (xx.x%) | xx (xx.x%) |
| Recent smoker | xx (xx.x%) | xx (xx.x%) | xx (xx.x%) |
| Previous smoker | xx (xx.x%) | xx (xx.x%) | xx (xx.x%) |

Note:
- (1) History of heavy drinking in the past (more than 5 days of excessive drinking within a month, more than 4 taels of liquor for men, more than 5 cans of beer; For women, more than 3 taels of baijiu and more than 4 cans of beer) - (2) Current smoking defined as smoking within 1 month (> 1 cigarette per day), recent smoking is defined as smoking between 1 month and 1 year (> 1 cigarette per day), previous smoking involves smoking for at least 1 year (smoking > 1 cigarette per day for more than half a year)

**Table 4.** Medical History at baseline.

| Medical History | Intervention | Control | Total |
| --- | --- | --- | --- |
| **Myocardial infarction** | xx (xx.x%) | xx (xx.x%) | xx (xx.x%) |
| Time since most recent episode (years) | N Mean(SE) | N Mean(SE) | N Mean(SE) |
| **Previous coronary stent implantation** | xx (xx.x%) | xx (xx.x%) | xx (xx.x%) |
| Time since most recent placement (years) | N Mean(SE) | N Mean(SE) | N Mean(SE) |
| **Previous CABG** | xx (xx.x%) | xx (xx.x%) | xx (xx.x%) |
| Time since most recent procedure (years) | N Mean(SE) | N Mean(SE) | N Mean(SE) |
| **Other confirmed coronary heart disease** | xx (xx.x%) | xx (xx.x%) | xx (xx.x%) |
| Coronary artery stenosis ≥ 50% (angiography finding) | xx (xx.x%) | xx (xx.x%) | xx (xx.x%) |
| Positive stress test and specific manifestation on imaging exam | xx (xx.x%) | xx (xx.x%) | xx (xx.x%) |
| Others |  |  |  |
| **Hypertension** | xx (xx.x%) | xx (xx.x%) | xx (xx.x%) |
| Time since first diagnosis | xx (xx.x%) | xx (xx.x%) | xx (xx.x%) |
| Unknown | xx (xx.x%) | xx (xx.x%) | xx (xx.x%) |
| **Diabetes** | xx (xx.x%) | xx (xx.x%) | xx (xx.x%) |
| Time since first diagnosis | xx (xx.x%) | xx (xx.x%) | xx (xx.x%) |
| Unknown | xx (xx.x%) | xx (xx.x%) | xx (xx.x%) |
| **Hyperlipidemia** | xx (xx.x%) | xx (xx.x%) | xx (xx.x%) |
| **Cardiac arrest, ventricular fibrillation or ventricular tachycardia with hemodynamic instability** | xx (xx.x%) | xx (xx.x%) | xx (xx.x%) |
| **Atrial fibrillation or flutter** | xx (xx.x%) | xx (xx.x%) | xx (xx.x%) |
| Time since diagnosis | N Mean(SE) | N Mean(SE) | N Mean(SE) |
| Paroxysmal | xx (xx.x%) | xx (xx.x%) | xx (xx.x%) |
| Persistent | xx (xx.x%) | xx (xx.x%) | xx (xx.x%) |
| **Congenital heart disease** | xx (xx.x%) | xx (xx.x%) | xx (xx.x%) |
| **History of pacemaker implantation** | xx (xx.x%) | xx (xx.x%) | xx (xx.x%) |
| **History of single or dual chamber pacemaker implantation** | xx (xx.x%) | xx (xx.x%) | xx (xx.x%) |
| **History of CRT implantation** | xx (xx.x%) | xx (xx.x%) | xx (xx.x%) |
| **History of ICD implantation** | xx (xx.x%) | xx (xx.x%) | xx (xx.x%) |
| **Previous cardiotoxic substance use** | xx (xx.x%) | xx (xx.x%) | xx (xx.x%) |
| Alcohol abuse for at least 5 years | xx (xx.x%) | xx (xx.x%) | xx (xx.x%) |
| Drug use | xx (xx.x%) | xx (xx.x%) | xx (xx.x%) |
| Other | xx (xx.x%) | xx (xx.x%) | xx (xx.x%) |
| **Cerebrovascular disease** | xx (xx.x%) | xx (xx.x%) | xx (xx.x%) |
| Cerebrovascular ischemic stroke | xx (xx.x%) | xx (xx.x%) | xx (xx.x%) |
| Hemorrhagic stroke | xx (xx.x%) | xx (xx.x%) | xx (xx.x%) |
| Transient ischemic attack (TIA) | xx (xx.x%) | xx (xx.x%) | xx (xx.x%) |
| Other | xx (xx.x%) | xx (xx.x%) | xx (xx.x%) |
| **Respiratory diseases** | xx (xx.x%) | xx (xx.x%) | xx (xx.x%) |
| Chronic obstructive pulmonary disease | xx (xx.x%) | xx (xx.x%) | xx (xx.x%) |
| Sleep apnea syndrome | xx (xx.x%) | xx (xx.x%) | xx (xx.x%) |
| Bronchial asthma | xx (xx.x%) | xx (xx.x%) | xx (xx.x%) |
| Pulmonary embolism | xx (xx.x%) | xx (xx.x%) | xx (xx.x%) |
| **Malignant tumors** | xx (xx.x%) | xx (xx.x%) | xx (xx.x%) |
| History of chemotherapy medication use | xx (xx.x%) | xx (xx.x%) | xx (xx.x%) |
| History of radiotherapy to the mediastinum | xx (xx.x%) | xx (xx.x%) | xx (xx.x%) |
| **Mental illness** | xx (xx.x%) | xx (xx.x%) | xx (xx.x%) |

**Table 5.** Clinical assessment at baseline.

|  | Intervention | Control | Total |
| --- | --- | --- | --- |
|  | N Mean (SD) | N Mean (SD) | N Mean (SD) |
| **Weight (kg)** |  |  |  |
| N Mean(SD) | N xx.x (xx.x) | N xx.x (xx.x) | N xx.x (xx.x) |
| Median [Q1 – Q3] | xx.x [xx.x- xx.x] | xx.x [xx.x- xx.x] | xx.x [xx.x- xx.x] |
| **BMI (kg/m<sup>2</sup>)** |  |  |  |
| N Mean(SD) | N xx.x (xx.x) | N xx.x (xx.x) | N xx.x (xx.x) |
| Median [Q1 – Q3] | xx.x [xx.x- xx.x] | xx.x [xx.x- xx.x] | xx.x [xx.x- xx.x] |
| **Systolic blood pressure (mmHg)** | N xx.x (xx.x) | N xx.x (xx.x) | N xx.x (xx.x) |
| N Mean(SD) | N xx.x (xx.x) | N xx.x (xx.x) | N xx.x (xx.x) |
| Median [Q1 – Q3] | xx.x [xx.x- xx.x] | xx.x [xx.x- xx.x] | xx.x [xx.x- xx.x] |
| **Diastolic blood pressure (mmHg)** | N xx.x (xx.x) | N xx.x (xx.x) | N xx.x (xx.x) |
| N Mean(SD) | N xx.x (xx.x) | N xx.x (xx.x) | N xx.x (xx.x) |
| Median [Q1 – Q3] | xx.x [xx.x- xx.x] | xx.x [xx.x- xx.x] | xx.x [xx.x- xx.x] |
| **Heart rate (bpm)** | N xx.x (xx.x) | N xx.x (xx.x) | N xx.x (xx.x) |
| **Time since first diagnosis of heart failure** | N xx.x (xx.x) | N xx.x (xx.x) | N xx.x (xx.x) |
| Unknown / Missing | xx (xx.x%) | xx (xx.x%) | xx (xx.x%) |
| **New York Heart Association NYHA functional class** |  |  |  |
| Level III | xx (xx.x%) | xx (xx.x%) | xx (xx.x%) |
| Level IV | xx (xx.x%) | xx (xx.x%) | xx (xx.x%) |

**Table 6.**
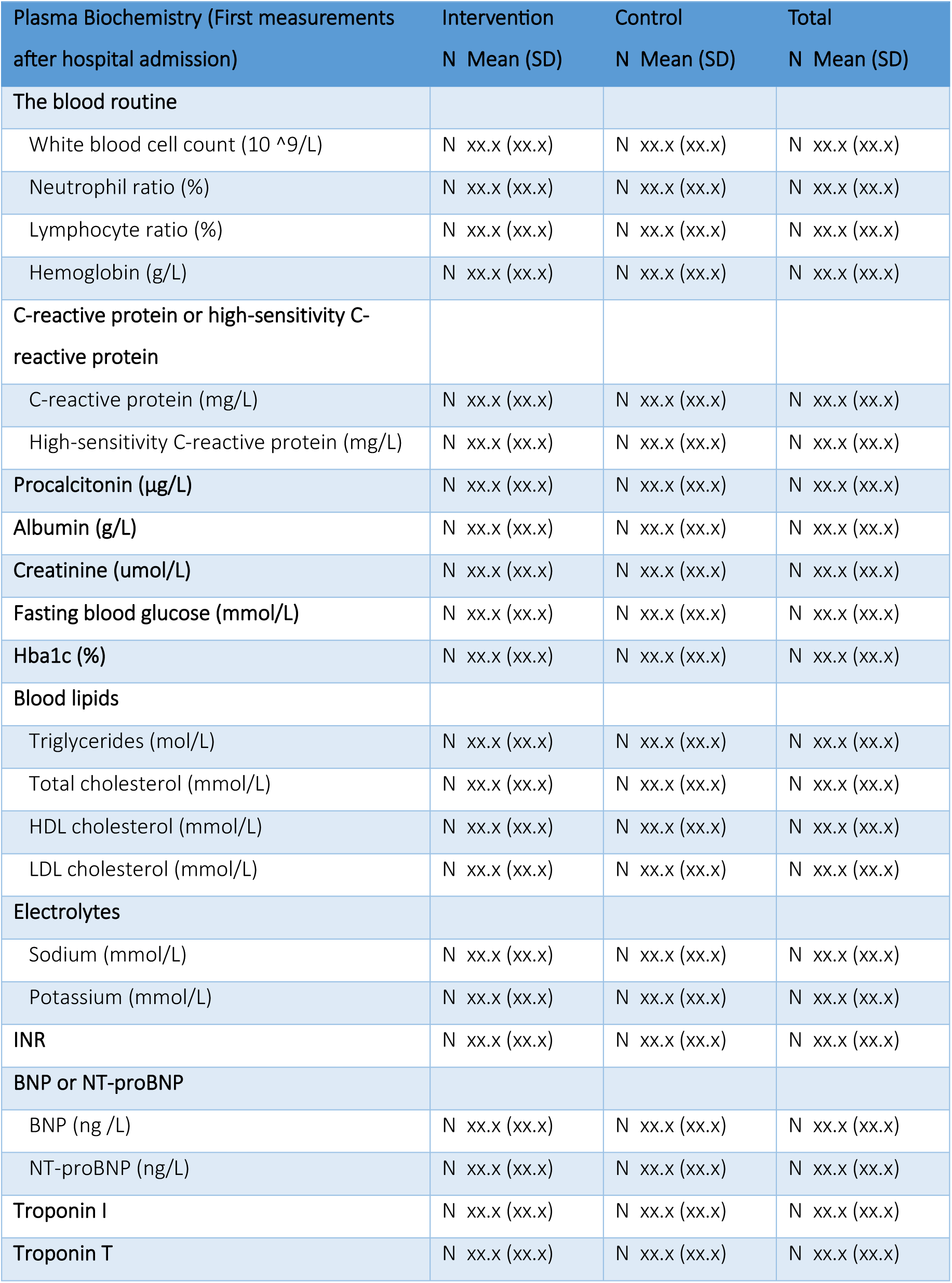
Laboratory measurements at baseline.

| Plasma Biochemistry (First measurements after hospital admission) | Intervention<br>N Mean (SD) | Control<br>N Mean (SD) | Total<br>N Mean (SD) |
| --- | --- | --- | --- |
| **The blood routine** |  |  |  |
| White blood cell count ($10^9/L$) | N xx.x (xx.x) | N xx.x (xx.x) | N xx.x (xx.x) |
| Neutrophil ratio (%) | N xx.x (xx.x) | N xx.x (xx.x) | N xx.x (xx.x) |
| Lymphocyte ratio (%) | N xx.x (xx.x) | N xx.x (xx.x) | N xx.x (xx.x) |
| Hemoglobin (g/L) | N xx.x (xx.x) | N xx.x (xx.x) | N xx.x (xx.x) |
| **C-reactive protein or high-sensitivity C-reactive protein** |  |  |  |
| C-reactive protein (mg/L) | N xx.x (xx.x) | N xx.x (xx.x) | N xx.x (xx.x) |
| High-sensitivity C-reactive protein (mg/L) | N xx.x (xx.x) | N xx.x (xx.x) | N xx.x (xx.x) |
| **Procalcitonin ($\mu g/L$)** | N xx.x (xx.x) | N xx.x (xx.x) | N xx.x (xx.x) |
| **Albumin (g/L)** | N xx.x (xx.x) | N xx.x (xx.x) | N xx.x (xx.x) |
| **Creatinine ($\mu mol/L$)** | N xx.x (xx.x) | N xx.x (xx.x) | N xx.x (xx.x) |
| **Fasting blood glucose (mmol/L)** | N xx.x (xx.x) | N xx.x (xx.x) | N xx.x (xx.x) |
| **Hba1c (%)** | N xx.x (xx.x) | N xx.x (xx.x) | N xx.x (xx.x) |
| **Blood lipids** |  |  |  |
| Triglycerides (mol/L) | N xx.x (xx.x) | N xx.x (xx.x) | N xx.x (xx.x) |
| Total cholesterol (mmol/L) | N xx.x (xx.x) | N xx.x (xx.x) | N xx.x (xx.x) |
| HDL cholesterol (mmol/L) | N xx.x (xx.x) | N xx.x (xx.x) | N xx.x (xx.x) |
| LDL cholesterol (mmol/L) | N xx.x (xx.x) | N xx.x (xx.x) | N xx.x (xx.x) |
| **Electrolytes** |  |  |  |
| Sodium (mmol/L) | N xx.x (xx.x) | N xx.x (xx.x) | N xx.x (xx.x) |
| Potassium (mmol/L) | N xx.x (xx.x) | N xx.x (xx.x) | N xx.x (xx.x) |
| **INR** | N xx.x (xx.x) | N xx.x (xx.x) | N xx.x (xx.x) |
| **BNP or NT-proBNP** |  |  |  |
| BNP (ng /L) | N xx.x (xx.x) | N xx.x (xx.x) | N xx.x (xx.x) |
| NT-proBNP (ng/L) | N xx.x (xx.x) | N xx.x (xx.x) | N xx.x (xx.x) |
| **Troponin I** | N xx.x (xx.x) | N xx.x (xx.x) | N xx.x (xx.x) |
| **Troponin T** | N xx.x (xx.x) | N xx.x (xx.x) | N xx.x (xx.x) |

**Table 7.** Diagnostic findings / methods at admission.

|  | Intervention | Control | Total |
| --- | --- | --- | --- |
|  | N Mean (SD) | N Mean (SD) | N Mean (SD) |
| **Echocardiography** |  |  |  |
| The left ventricular ejection fraction (EF) (%) | N xx.x (xx.x) | N xx.x (xx.x) | N xx.x (xx.x) |
| **Other valve problems** |  |  |  |
| **Aortic stenosis** | xx (xx.x%) | xx (xx.x%) | xx (xx.x%) |
| Mild | xx (xx.x%) | xx (xx.x%) | xx (xx.x%) |
| Moderate | xx (xx.x%) | xx (xx.x%) | xx (xx.x%) |
| Severe | xx (xx.x%) | xx (xx.x%) | xx (xx.x%) |
| **Aortic insufficiency** | xx (xx.x%) | xx (xx.x%) | xx (xx.x%) |
| Mild | xx (xx.x%) | xx (xx.x%) | xx (xx.x%) |
| Moderate | xx (xx.x%) | xx (xx.x%) | xx (xx.x%) |
| Severe | xx (xx.x%) | xx (xx.x%) | xx (xx.x%) |
| **Mitral regurgitation** | xx (xx.x%) | xx (xx.x%) | xx (xx.x%) |
| Mild | xx (xx.x%) | xx (xx.x%) | xx (xx.x%) |
| Moderate | xx (xx.x%) | xx (xx.x%) | xx (xx.x%) |
| Severe | xx (xx.x%) | xx (xx.x%) | xx (xx.x%) |
| **Mitral stenosis** | xx (xx.x%) | xx (xx.x%) | xx (xx.x%) |
| Mild | xx (xx.x%) | xx (xx.x%) | xx (xx.x%) |
| Moderate | xx (xx.x%) | xx (xx.x%) | xx (xx.x%) |
| Severe | xx (xx.x%) | xx (xx.x%) | xx (xx.x%) |
| **Tricuspid insufficiency** | xx (xx.x%) | xx (xx.x%) | xx (xx.x%) |
| Mild | xx (xx.x%) | xx (xx.x%) | xx (xx.x%) |
| Moderate | xx (xx.x%) | xx (xx.x%) | xx (xx.x%) |
| Severe | xx (xx.x%) | xx (xx.x%) | xx (xx.x%) |
| **Other Diagnostic methods** |  |  |  |
| **Electrocardiogram** |  |  |  |
| Sinus rhythm | xx (xx.x%) | xx (xx.x%) | xx (xx.x%) |
| Left bundle branch block | xx (xx.x%) | xx (xx.x%) | xx (xx.x%) |
| The QRS duration (ms) | N xx.x (xx.x) | N xx.x (xx.x) | N xx.x (xx.x) |
| **Chest radiograph** | xx (xx.x%) | xx (xx.x%) | xx (xx.x%) |
| Pleural effusion | xx (xx.x%) | xx (xx.x%) | xx (xx.x%) |
| **Coronary angiography** | xx (xx.x%) | xx (xx.x%) | xx (xx.x%) |
| Presence of greater than 50% coronary stenosis | xx (xx.x%) | xx (xx.x%) | xx (xx.x%) |

**Table 8.** Medication / Interventional treatment in the hospital.

|  | Intervention<br>N Mean (SD) | Control<br>N Mean (SD) | Total<br>N Mean (SD) |
| --- | --- | --- | --- |
| **Medications in the hospital** |  |  |  |
| **Use of antibiotics in the hospital** | xx (xx.x%) | xx (xx.x%) | xx (xx.x%) |
| Duration of antibiotic use (days) | N Mean (SD) | N Mean (SD) | N Mean (SD) |
| **Intravenous inotropic drugs <sup>(1)</sup>** |  |  |  |
| **Intravenous vasodilators <sup>(2)</sup>** | xx (xx.x%) | xx (xx.x%) | xx (xx.x%) |
| **Total amount of infusion in the first 24 hours after admission (ml)** | N Mean (SD) | N Mean (SD) | N Mean (SD) |
| **Interventional treatment received during admission** |  |  |  |
| **Permanent pacemaker (single or dual chamber) insertion** | xx (xx.x%) | xx (xx.x%) | xx (xx.x%) |
| Time since surgery (days) | N Mean (SD) | N Mean (SD) | N Mean (SD) |
| **Implantable Cardioverter defibrillator (ICD)** | xx (xx.x%) | xx (xx.x%) | xx (xx.x%) |
| Time since surgery (days) | N Mean (SD) | N Mean (SD) | N Mean (SD) |
| **Cardiac resynchronization (CRT) implantation** | xx (xx.x%) | xx (xx.x%) | xx (xx.x%) |
| Time since surgery (days) | N Mean (SD) | N Mean (SD) | N Mean (SD) |
| **Percutaneous coronary intervention (PCI)** | xx (xx.x%) | xx (xx.x%) | xx (xx.x%) |
| Time since surgery (days) | N Mean (SD) | N Mean (SD) | N Mean (SD) |
| **Other percutaneous procedures** | xx (xx.x%) | xx (xx.x%) | xx (xx.x%) |
| Time since surgery (days) | N Mean (SD) | N Mean (SD) | N Mean (SD) |
| **Permanent pacemaker insertion (single/dual)** | xx (xx.x%) | xx (xx.x%) | xx (xx.x%) |
| Time since surgery (days) | N Mean (SD) | N Mean (SD) | N Mean (SD) |
| **Implantable cardioverter defibrillator (ICD)** | xx (xx.x%) | xx (xx.x%) | xx (xx.x%) |
| Time since surgery (days) | N Mean (SD) | N Mean (SD) | N Mean (SD) |

Note:
(1) Intravenous inotrope drugs incl. dopamine, dobutamine, levosimendan, cedilanid, milrinone
(2) Intravenous vasodilators incl. nitroglycerin, isosorbide mononitrate, sodium nitroprusside, nesiritide, urapidil

**Table 9.**
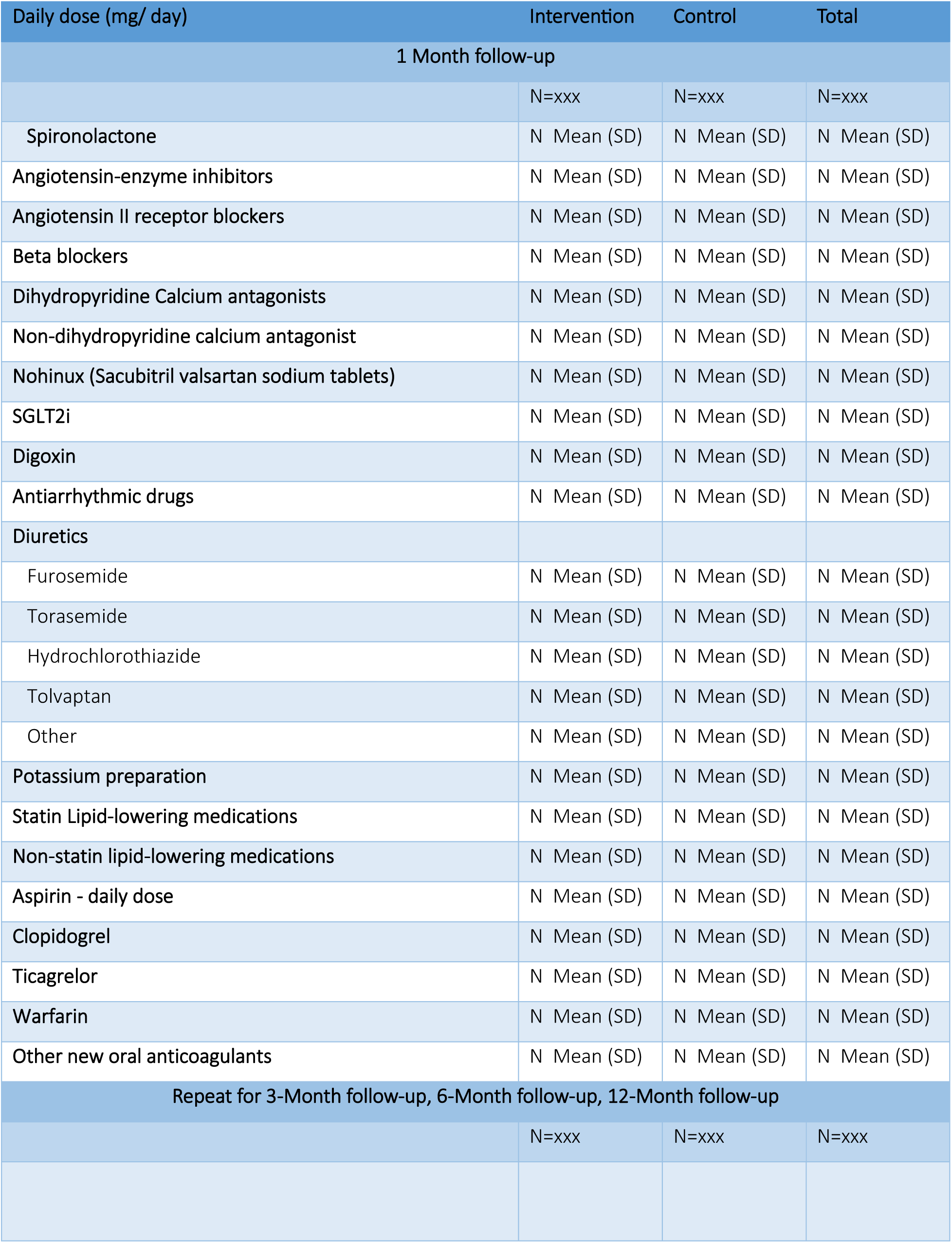
Summary medication dosage (taken since discharge) at follow-up timepoints.

**Table 10.** Summary detailed medication usage (taken since discharge) at follow-up timepoints.

|  | Intervention | Control | Total |
| --- | --- | --- | --- |
| **1 Month follow-up** |  |  |  |
|  | N=xxx | N=xxx | N=xxx |
| **Spironolactone** | xx (xx.x%) | xx (xx.x%) | xx (xx.x%) |
| Daily dose (mg/ day) | N Mean (SD) | N Mean (SD) | N Mean (SD) |
| **Angiotensin-enzyme inhibitors** | xx (xx.x%) | xx (xx.x%) | xx (xx.x%) |
| Daily dose (mg/ day) | N Mean (SD) | N Mean (SD) | N Mean (SD) |
| Captopril | xx (xx.x%) | xx (xx.x%) | xx (xx.x%) |
| Enalapril | xx (xx.x%) | xx (xx.x%) | xx (xx.x%) |
| Perindopril | xx (xx.x%) | xx (xx.x%) | xx (xx.x%) |
| Benazepril | xx (xx.x%) | xx (xx.x%) | xx (xx.x%) |
| Silapril | xx (xx.x%) | xx (xx.x%) | xx (xx.x%) |
| Ramipril | xx (xx.x%) | xx (xx.x%) | xx (xx.x%) |
| Fosinopril | xx (xx.x%) | xx (xx.x%) | xx (xx.x%) |
| Imidapril | xx (xx.x%) | xx (xx.x%) | xx (xx.x%) |
| Lisinopril | xx (xx.x%) | xx (xx.x%) | xx (xx.x%) |
| Other | xx (xx.x%) | xx (xx.x%) | xx (xx.x%) |
| **Angiotensin II receptor blockers** | xx (xx.x%) | xx (xx.x%) | xx (xx.x%) |
| Daily dose (mg/ day) | N Mean (SD) | N Mean (SD) | N Mean (SD) |
| Irbesartan | xx (xx.x%) | xx (xx.x%) | xx (xx.x%) |
| Valsartan | xx (xx.x%) | xx (xx.x%) | xx (xx.x%) |
| Telmisartan | xx (xx.x%) | xx (xx.x%) | xx (xx.x%) |
| Candesartan | xx (xx.x%) | xx (xx.x%) | xx (xx.x%) |
| Losartan | xx (xx.x%) | xx (xx.x%) | xx (xx.x%) |
| Epresartan | xx (xx.x%) | xx (xx.x%) | xx (xx.x%) |
| Olmesartan | xx (xx.x%) | xx (xx.x%) | xx (xx.x%) |
| Other | xx (xx.x%) | xx (xx.x%) | xx (xx.x%) |
| **Beta blockers** | xx (xx.x%) | xx (xx.x%) | xx (xx.x%) |
| Daily dose (mg/ day) | N Mean (SD) | N Mean (SD) | N Mean (SD) |
| Propranolol | xx (xx.x%) | xx (xx.x%) | xx (xx.x%) |
| Metoprolol | xx (xx.x%) | xx (xx.x%) | xx (xx.x%) |
| Metoprolol extended-release tablets | xx (xx.x%) | xx (xx.x%) | xx (xx.x%) |
| Bisoprolol | xx (xx.x%) | xx (xx.x%) | xx (xx.x%) |
| Carvedilol | xx (xx.x%) | xx (xx.x%) | xx (xx.x%) |
| Other | xx (xx.x%) | xx (xx.x%) | xx (xx.x%) |
| **Dihydropyridine Calcium antagonists** | xx (xx.x%) | xx (xx.x%) | xx (xx.x%) |
| Daily dose (mg/ day) | N Mean (SD) | N Mean (SD) | N Mean (SD) |
| Amlodipine | xx (xx.x%) | xx (xx.x%) | xx (xx.x%) |
| Nifedipine | xx (xx.x%) | xx (xx.x%) | xx (xx.x%) |
| Felodipine | xx (xx.x%) | xx (xx.x%) | xx (xx.x%) |
| Lacidipine | xx (xx.x%) | xx (xx.x%) | xx (xx.x%) |
| Benidipine | xx (xx.x%) | xx (xx.x%) | xx (xx.x%) |
| Nicardipine | xx (xx.x%) | xx (xx.x%) | xx (xx.x%) |
| Other | xx (xx.x%) | xx (xx.x%) | xx (xx.x%) |
| **Non-dihydropyridine calcium antagonist** | xx (xx.x%) | xx (xx.x%) | xx (xx.x%) |
| Daily dose (mg/ day) | N Mean (SD) | N Mean (SD) | N Mean (SD) |
| Verapamil | xx (xx.x%) | xx (xx.x%) | xx (xx.x%) |
| Diltiazem | xx (xx.x%) | xx (xx.x%) | xx (xx.x%) |
| Other | xx (xx.x%) | xx (xx.x%) | xx (xx.x%) |
| **Nohinux (Sacubitril valsartan sodium tablets)** | xx (xx.x%) | xx (xx.x%) | xx (xx.x%) |
| Daily dose (mg/ day) | N Mean (SD) | N Mean (SD) | N Mean (SD) |
| **SGLT2i** | xx (xx.x%) | xx (xx.x%) | xx (xx.x%) |
| Daily dose (mg/ day) | N Mean (SD) | N Mean (SD) | N Mean (SD) |
| Dapagliflozin | xx (xx.x%) | xx (xx.x%) | xx (xx.x%) |
| Canagliflozin | xx (xx.x%) | xx (xx.x%) | xx (xx.x%) |
| Other | xx (xx.x%) | xx (xx.x%) | xx (xx.x%) |
| **Digoxin** | xx (xx.x%) | xx (xx.x%) | xx (xx.x%) |
| Daily dose (mg/ day) | N Mean (SD) | N Mean (SD) | N Mean (SD) |
| **Antiarrhythmic drugs** | xx (xx.x%) | xx (xx.x%) | xx (xx.x%) |
| Daily dose (mg/ day) | N Mean (SD) | N Mean (SD) | N Mean (SD) |
| Propafenone | xx (xx.x%) | xx (xx.x%) | xx (xx.x%) |
| Amiodarone | xx (xx.x%) | xx (xx.x%) | xx (xx.x%) |
| Sotalol | xx (xx.x%) | xx (xx.x%) | xx (xx.x%) |
| Other | xx (xx.x%) | xx (xx.x%) | xx (xx.x%) |
| **Diuretics** | xx (xx.x%) | xx (xx.x%) | xx (xx.x%) |
| Furosemide - daily dose (mg/ day) | N Mean (SD) | N Mean (SD) | N Mean (SD) |
| Torsemide- daily dose (mg/ day) | N Mean (SD) | N Mean (SD) | N Mean (SD) |
| Hydrochlorothiazide - daily dose (mg/ day) | N Mean (SD) | N Mean (SD) | N Mean (SD) |
| Tolvaptan - daily dose (mg/ day) | N Mean (SD) | N Mean (SD) | N Mean (SD) |
| Other - daily dose (mg/ day) | N Mean (SD) | N Mean (SD) | N Mean (SD) |
| **Potassium preparation** | xx (xx.x%) | xx (xx.x%) | xx (xx.x%) |
| Daily dose (mg/ day) | N Mean (SD) | N Mean (SD) | N Mean (SD) |
| Potassium chloride tablets | xx (xx.x%) | xx (xx.x%) | xx (xx.x%) |
| Potassium citrate | xx (xx.x%) | xx (xx.x%) | xx (xx.x%) |
| Potassium magnesium aspartate tablets | xx (xx.x%) | xx (xx.x%) | xx (xx.x%) |
| Other | xx (xx.x%) | xx (xx.x%) | xx (xx.x%) |
| **Statin Lipid-lowering medications** | xx (xx.x%) | xx (xx.x%) | xx (xx.x%) |
| Daily dose (mg/ day) | N Mean (SD) | N Mean (SD) | N Mean (SD) |
| Atorvastatin | xx (xx.x%) | xx (xx.x%) | xx (xx.x%) |
| Rosuvastatin | xx (xx.x%) | xx (xx.x%) | xx (xx.x%) |
| Fluvastatin | xx (xx.x%) | xx (xx.x%) | xx (xx.x%) |
| Pitavastatin | xx (xx.x%) | xx (xx.x%) | xx (xx.x%) |
| Simvastatin | xx (xx.x%) | xx (xx.x%) | xx (xx.x%) |
| Ovastatin | xx (xx.x%) | xx (xx.x%) | xx (xx.x%) |
| Pravastatin | xx (xx.x%) | xx (xx.x%) | xx (xx.x%) |
| Other | xx (xx.x%) | xx (xx.x%) | xx (xx.x%) |
| **Non-statin lipid-lowering medications** | xx (xx.x%) | xx (xx.x%) | xx (xx.x%) |
| Daily dose (mg/ day) | N Mean (SD) | N Mean (SD) | N Mean (SD) |
| Ezetimibe | xx (xx.x%) | xx (xx.x%) | xx (xx.x%) |
| other | xx (xx.x%) | xx (xx.x%) | xx (xx.x%) |
| **Aspirin - daily dose (mg/ day)** | xx (xx.x%) | xx (xx.x%) | xx (xx.x%) |
| Daily dose (mg/ day) | N Mean (SD) | N Mean (SD) | N Mean (SD) |
| **Clopidogrel - daily dose (mg/ day)** | xx (xx.x%) | xx (xx.x%) | xx (xx.x%) |
| Daily dose (mg/ day) | N Mean (SD) | N Mean (SD) | N Mean (SD) |
| **Ticagrelor - daily dose (mg/ day)** | xx (xx.x%) | xx (xx.x%) | xx (xx.x%) |
| Daily dose (mg/ day) | N Mean (SD) | N Mean (SD) | N Mean (SD) |
| **Warfarin - daily dose (mg/ day)** | xx (xx.x%) | xx (xx.x%) | xx (xx.x%) |
| Daily dose (mg/ day) | N Mean (SD) | N Mean (SD) | N Mean (SD) |
| **Other new oral anticoagulants - daily dose (mg/ day)** | xx (xx.x%) | xx (xx.x%) | xx (xx.x%) |
| Daily dose (mg/ day) | N Mean (SD) | N Mean (SD) | N Mean (SD) |
| Dabigatran | xx (xx.x%) | xx (xx.x%) | xx (xx.x%) |
| Rivaroxaban | xx (xx.x%) | xx (xx.x%) | xx (xx.x%) |
| Edoxaban | xx (xx.x%) | xx (xx.x%) | xx (xx.x%) |
| Apixaban | xx (xx.x%) | xx (xx.x%) | xx (xx.x%) |
| Other | xx (xx.x%) | xx (xx.x%) | xx (xx.x%) |
| Repeat for 3-Month follow-up, 6-Month follow-up, 12-Month follow-up |  |  |  |
|  | N=xxx | N=xxx | N=xxx |

**Table 11.**
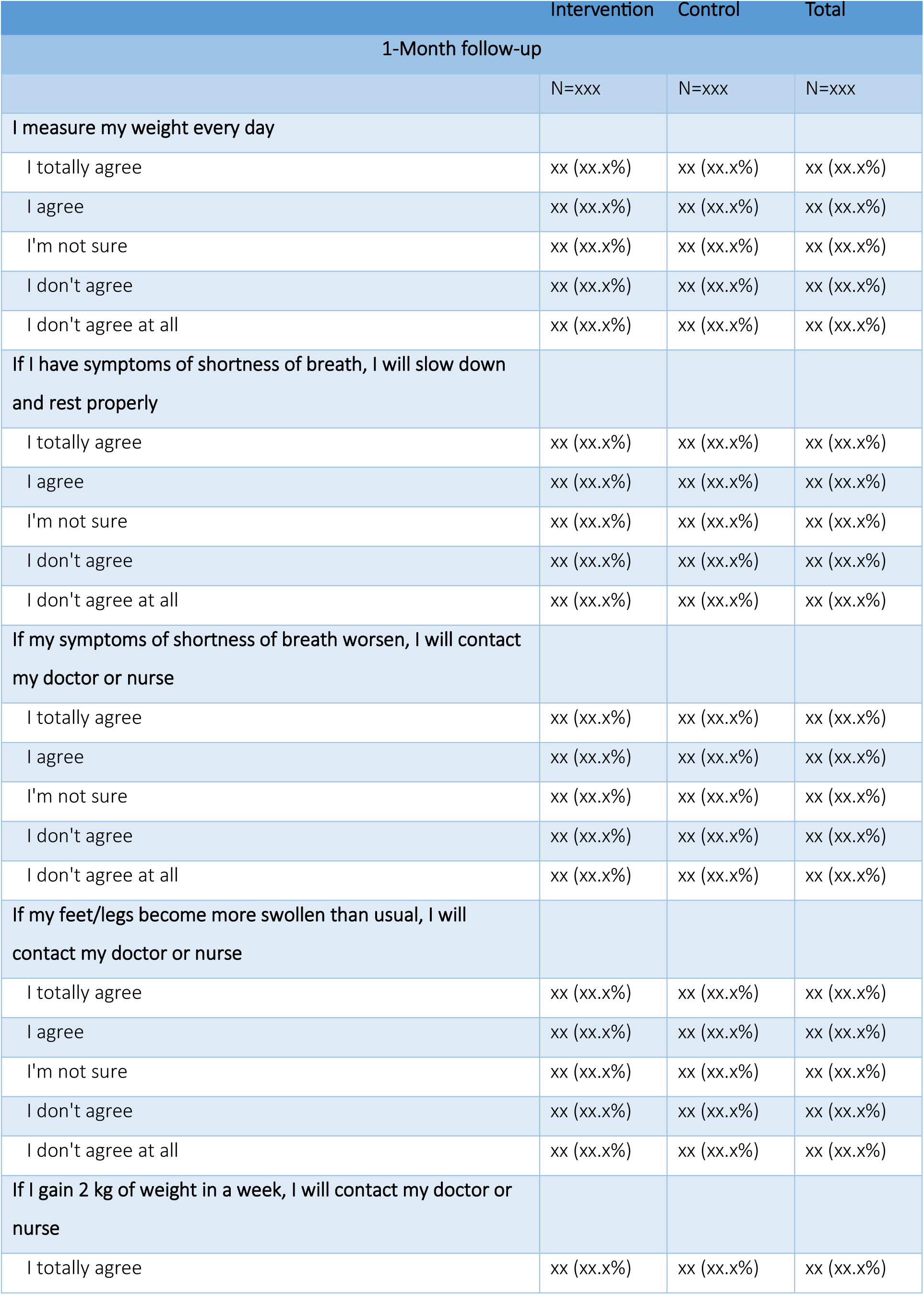
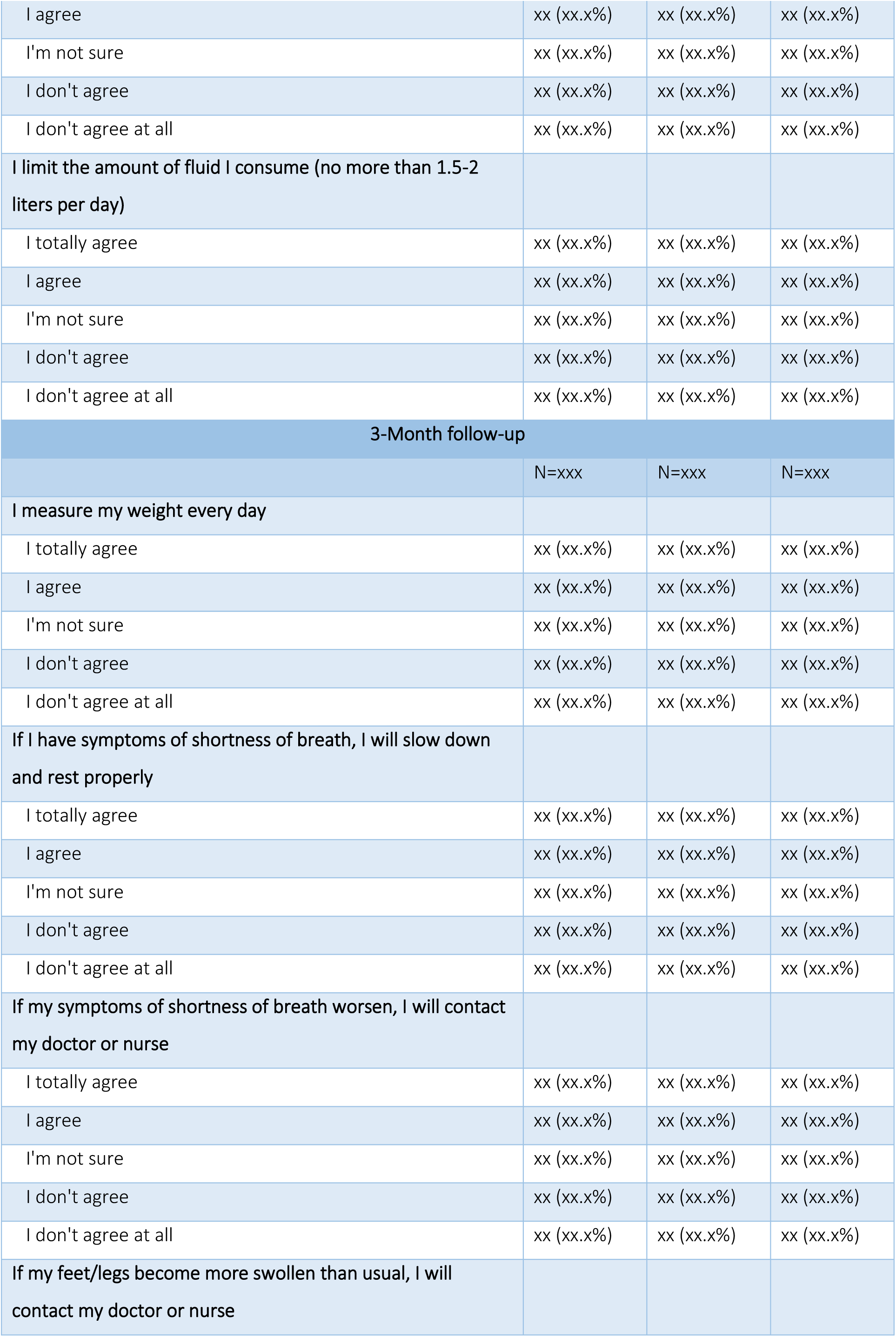
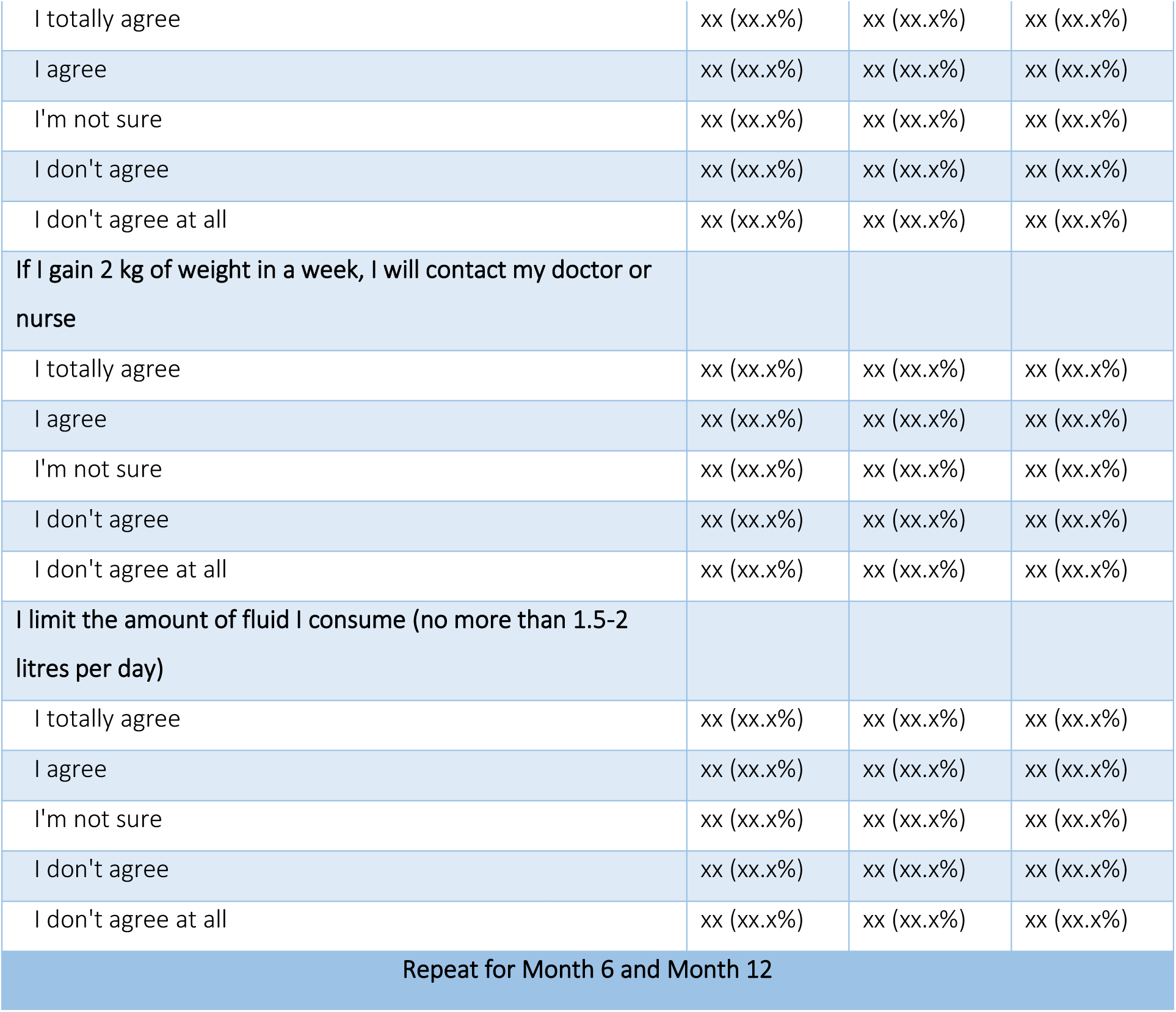
Heart failure self-management scale – descriptive table by timepoints (Month 1, 3, 6, 12)

**Table 12.** EQ5D-5L – descriptive table by timepoints.

|  | Intervention | Control | Total |
| --- | --- | --- | --- |
| **Baseline** |  |  |  |
|  | N=xxx | N=xxx | N=xxx |
| **Mobility** |  |  |  |
| I have no trouble getting around | xx (xx.x%) | xx (xx.x%) | xx (xx.x%) |
| I have some difficulty getting around | xx (xx.x%) | xx (xx.x%) | xx (xx.x%) |
| I can't get out of bed and walk around | xx (xx.x%) | xx (xx.x%) | xx (xx.x%) |
| **Self-care ability** |  |  |  |
| I have no difficulty taking care of myself | xx (xx.x%) | xx (xx.x%) | xx (xx.x%) |
| I have some difficulty bathing or dressing myself | xx (xx.x%) | xx (xx.x%) | xx (xx.x%) |
| I can't bathe or dress myself | xx (xx.x%) | xx (xx.x%) | xx (xx.x%) |
| **Ability to perform daily activities (such as work, school, housework, family or leisure activities)** |  |  |  |
| I have no difficulty performing daily activities | xx (xx.x%) | xx (xx.x%) | xx (xx.x%) |
| I have some difficulty with my daily activities | xx (xx.x%) | xx (xx.x%) | xx (xx.x%) |
| I can't carry out my daily activities | xx (xx.x%) | xx (xx.x%) | xx (xx.x%) |
| **Pain or discomfort** |  |  |  |
| I don't have any pain or discomfort | xx (xx.x%) | xx (xx.x%) | xx (xx.x%) |
| I had moderate pain or discomfort | xx (xx.x%) | xx (xx.x%) | xx (xx.x%) |
| I have severe pain or discomfort | xx (xx.x%) | xx (xx.x%) | xx (xx.x%) |
| **Anxiety or depression** |  |  |  |
| I don't have any anxiety or depression | xx (xx.x%) | xx (xx.x%) | xx (xx.x%) |
| I have moderate anxiety or depression | xx (xx.x%) | xx (xx.x%) | xx (xx.x%) |
| I have severe anxiety or depression | xx (xx.x%) | xx (xx.x%) | xx (xx.x%) |
| **Health utility score – mean(SD)** | xx.xx (xx.x) | xx.xx (xx.x) | xx.xx (xx.x) |
| **1 Month follow-up** |  |  |  |
|  | N=xxx | N=xxx | N=xxx |
| **Mobility** |  |  |  |
| I have no trouble getting around | xx (xx.x%) | xx (xx.x%) | xx (xx.x%) |
| I have some difficulty getting around | xx (xx.x%) | xx (xx.x%) | xx (xx.x%) |
| I can't get out of bed and walk around | xx (xx.x%) | xx (xx.x%) | xx (xx.x%) |
| **Self-care ability** |  |  |  |
| I have no difficulty taking care of myself | xx (xx.x%) | xx (xx.x%) | xx (xx.x%) |
| I have some difficulty bathing or dressing myself | xx (xx.x%) | xx (xx.x%) | xx (xx.x%) |
| I can't bathe or dress myself | xx (xx.x%) | xx (xx.x%) | xx (xx.x%) |
| **Ability to perform daily activities (such as work, school, housework, family or leisure activities)** |  |  |  |
| I have no difficulty performing daily activities | xx (xx.x%) | xx (xx.x%) | xx (xx.x%) |
| I have some difficulty with my daily activities | xx (xx.x%) | xx (xx.x%) | xx (xx.x%) |
| I can't carry out my daily activities | xx (xx.x%) | xx (xx.x%) | xx (xx.x%) |
| **Pain or discomfort** |  |  |  |
| I don't have any pain or discomfort | xx (xx.x%) | xx (xx.x%) | xx (xx.x%) |
| I had moderate pain or discomfort | xx (xx.x%) | xx (xx.x%) | xx (xx.x%) |
| I have severe pain or discomfort | xx (xx.x%) | xx (xx.x%) | xx (xx.x%) |
| **Anxiety or depression** |  |  |  |
| I don't have any anxiety or depression | xx (xx.x%) | xx (xx.x%) | xx (xx.x%) |
| I have moderate anxiety or depression | xx (xx.x%) | xx (xx.x%) | xx (xx.x%) |
| I have severe anxiety or depression | xx (xx.x%) | xx (xx.x%) | xx (xx.x%) |
| **Health utility score – mean(SD)** | xx.xx (xx.x) | xx.xx (xx.x) | xx.xx (xx.x) |
| **3 Months follow-up** |  |  |  |
|  | N=xxx | N=xxx | N=xxx |
| **Mobility** |  |  |  |
| I have no trouble getting around | xx (xx.x%) | xx (xx.x%) | xx (xx.x%) |
| I have some difficulty getting around | xx (xx.x%) | xx (xx.x%) | xx (xx.x%) |
| I can't get out of bed and walk around | xx (xx.x%) | xx (xx.x%) | xx (xx.x%) |
| **Self-care ability** |  |  |  |
| I have no difficulty taking care of myself | xx (xx.x%) | xx (xx.x%) | xx (xx.x%) |
| I have some difficulty bathing or dressing myself | xx (xx.x%) | xx (xx.x%) | xx (xx.x%) |
| I can't bathe or dress myself | xx (xx.x%) | xx (xx.x%) | xx (xx.x%) |
| **Ability to perform daily activities (such as work, school, housework, family or leisure activities)** |  |  |  |
| I have no difficulty performing daily activities | xx (xx.x%) | xx (xx.x%) | xx (xx.x%) |
| I have some difficulty with my daily activities | xx (xx.x%) | xx (xx.x%) | xx (xx.x%) |
| I can't carry out my daily activities | xx (xx.x%) | xx (xx.x%) | xx (xx.x%) |
| **Pain or discomfort** |  |  |  |
| I don't have any pain or discomfort | xx (xx.x%) | xx (xx.x%) | xx (xx.x%) |
| I had moderate pain or discomfort | xx (xx.x%) | xx (xx.x%) | xx (xx.x%) |
| I have severe pain or discomfort | xx (xx.x%) | xx (xx.x%) | xx (xx.x%) |
| **Anxiety or depression** |  |  |  |
| I don't have any anxiety or depression | xx (xx.x%) | xx (xx.x%) | xx (xx.x%) |
| I have moderate anxiety or depression | xx (xx.x%) | xx (xx.x%) | xx (xx.x%) |
| I have severe anxiety or depression | xx (xx.x%) | xx (xx.x%) | xx (xx.x%) |
| **Health utility score – mean(SD)** | xx.xx (xx.x) | xx.xx (xx.x) | xx.xx (xx.x) |
| **6 Months follow-up** |  |  |  |
|  | N=xxx | N=xxx | N=xxx |
| **Mobility** |  |  |  |
| I have no trouble getting around | xx (xx.x%) | xx (xx.x%) | xx (xx.x%) |
| I have some difficulty getting around | xx (xx.x%) | xx (xx.x%) | xx (xx.x%) |
| I can't get out of bed and walk around | xx (xx.x%) | xx (xx.x%) | xx (xx.x%) |
| **Self-care ability** |  |  |  |
| I have no difficulty taking care of myself | xx (xx.x%) | xx (xx.x%) | xx (xx.x%) |
| I have some difficulty bathing or dressing myself | xx (xx.x%) | xx (xx.x%) | xx (xx.x%) |
| I can't bathe or dress myself | xx (xx.x%) | xx (xx.x%) | xx (xx.x%) |
| **Ability to perform daily activities (such as work, school, housework, family or leisure activities)** |  |  |  |
| I have no difficulty performing daily activities | xx (xx.x%) | xx (xx.x%) | xx (xx.x%) |
| I have some difficulty with my daily activities | xx (xx.x%) | xx (xx.x%) | xx (xx.x%) |
| I can't carry out my daily activities | xx (xx.x%) | xx (xx.x%) | xx (xx.x%) |
| **Pain or discomfort** |  |  |  |
| I don't have any pain or discomfort | xx (xx.x%) | xx (xx.x%) | xx (xx.x%) |
| I had moderate pain or discomfort | xx (xx.x%) | xx (xx.x%) | xx (xx.x%) |
| I have severe pain or discomfort | xx (xx.x%) | xx (xx.x%) | xx (xx.x%) |
| **Anxiety or depression** |  |  |  |
| I don't have any anxiety or depression | xx (xx.x%) | xx (xx.x%) | xx (xx.x%) |
| I have moderate anxiety or depression | xx (xx.x%) | xx (xx.x%) | xx (xx.x%) |
| I have severe anxiety or depression | xx (xx.x%) | xx (xx.x%) | xx (xx.x%) |
| **Health utility score – mean(SD)** | xx.xx (xx.x) | xx.xx (xx.x) | xx.xx (xx.x) |
| **12 Months follow-up** |  |  |  |
|  | N=xxx | N=xxx | N=xxx |
| **Mobility** |  |  |  |
| I have no trouble getting around | xx (xx.x%) | xx (xx.x%) | xx (xx.x%) |
| I have some difficulty getting around | xx (xx.x%) | xx (xx.x%) | xx (xx.x%) |
| I can't get out of bed and walk around | xx (xx.x%) | xx (xx.x%) | xx (xx.x%) |
| **Self-care ability** |  |  |  |
| I have no difficulty taking care of myself | xx (xx.x%) | xx (xx.x%) | xx (xx.x%) |
| I have some difficulty bathing or dressing myself | xx (xx.x%) | xx (xx.x%) | xx (xx.x%) |
| I can't bathe or dress myself | xx (xx.x%) | xx (xx.x%) | xx (xx.x%) |
| **Ability to perform daily activities (such as work, school, housework, family or leisure activities)** |  |  |  |
| I have no difficulty performing daily activities | xx (xx.x%) | xx (xx.x%) | xx (xx.x%) |
| I have some difficulty with my daily activities | xx (xx.x%) | xx (xx.x%) | xx (xx.x%) |
| I can't carry out my daily activities | xx (xx.x%) | xx (xx.x%) | xx (xx.x%) |
| **Pain or discomfort** |  |  |  |
| I don't have any pain or discomfort | xx (xx.x%) | xx (xx.x%) | xx (xx.x%) |
| I had moderate pain or discomfort | xx (xx.x%) | xx (xx.x%) | xx (xx.x%) |
| I have severe pain or discomfort | xx (xx.x%) | xx (xx.x%) | xx (xx.x%) |
| **Anxiety or depression** |  |  |  |
| I don't have any anxiety or depression | xx (xx.x%) | xx (xx.x%) | xx (xx.x%) |
| I have moderate anxiety or depression | xx (xx.x%) | xx (xx.x%) | xx (xx.x%) |
| I have severe anxiety or depression | xx (xx.x%) | xx (xx.x%) | xx (xx.x%) |
| **Health utility score – mean(SD)** | xx.xx (xx.x) | xx.xx (xx.x) | xx.xx (xx.x) |

**Table 13.** Follow-up assessment of influenza-like infection symptoms.

| Timepoints | Intervention | Control | Total |
| --- | --- | --- | --- |
| **1-Month follow-up** |  |  |  |
|  | N=xxx | N=xxx | N=xxx |
| **Method of follow-up** |  |  |  |
| By telephone | xx (xx.x%) | xx (xx.x%) | xx (xx.x%) |
| By outpatient | xx (xx.x%) | xx (xx.x%) | xx (xx.x%) |
| **Influenza-like symptoms since discharge** |  |  |  |
| Fever $\geq 38^{\circ}$ C | xx (xx.x%) | xx (xx.x%) | xx (xx.x%) |
| Cough | xx (xx.x%) | xx (xx.x%) | xx (xx.x%) |
| Sore throat | xx (xx.x%) | xx (xx.x%) | xx (xx.x%) |
| If you received the influenza vaccine, whether you have had an adverse vaccination reaction within 7 days of vaccination | xx (xx.x%) | xx (xx.x%) | xx (xx.x%) |
| **3-Month follow-up** |  |  |  |
|  | N=xxx | N=xxx | N=xxx |
| By telephone | xx (xx.x%) | xx (xx.x%) | xx (xx.x%) |
| By outpatient | xx (xx.x%) | xx (xx.x%) | xx (xx.x%) |
| **Influenza-like symptoms since discharge** |  |  |  |
| Fever $\geq 38^{\circ}$ C | xx (xx.x%) | xx (xx.x%) | xx (xx.x%) |
| Cough | xx (xx.x%) | xx (xx.x%) | xx (xx.x%) |
| Sore throat | xx (xx.x%) | xx (xx.x%) | xx (xx.x%) |
| If you received the influenza vaccine, whether you have had an adverse vaccination reaction within 7 days of vaccination | xx (xx.x%) | xx (xx.x%) | xx (xx.x%) |
| **6-Month follow-up** |  |  |  |
|  | N=xxx | N=xxx | N=xxx |
| **Method of follow-up** |  |  |  |
| By telephone | xx (xx.x%) | xx (xx.x%) | xx (xx.x%) |
| By outpatient | xx (xx.x%) | xx (xx.x%) | xx (xx.x%) |
| **Influenza-like symptoms since discharge** |  |  |  |
| Fever $\geq 38^{\circ}$ C | xx (xx.x%) | xx (xx.x%) | xx (xx.x%) |
| Cough | xx (xx.x%) | xx (xx.x%) | xx (xx.x%) |
| Sore throat | xx (xx.x%) | xx (xx.x%) | xx (xx.x%) |
| If you received the influenza vaccine, whether you have had an adverse vaccination reaction within 7 days of vaccination | xx (xx.x%) | xx (xx.x%) | xx (xx.x%) |
| **12-Month follow-up** |  |  |  |
|  | N=xxx | N=xxx | N=xxx |
| **Method of follow-up** |  |  |  |
| By telephone | xx (xx.x%) | xx (xx.x%) | xx (xx.x%) |
| By outpatient | xx (xx.x%) | xx (xx.x%) | xx (xx.x%) |
| **Influenza-like symptoms since discharge** |  |  |  |
| Fever $\geq 38^{\circ}$ C | xx (xx.x%) | xx (xx.x%) | xx (xx.x%) |
| Cough | xx (xx.x%) | xx (xx.x%) | xx (xx.x%) |
| Sore throat | xx (xx.x%) | xx (xx.x%) | xx (xx.x%) |
| If you received the influenza vaccine, whether you have had an adverse vaccination reaction within 7 days of vaccination | xx (xx.x%) | xx (xx.x%) | xx (xx.x%) |

**Table 14.**
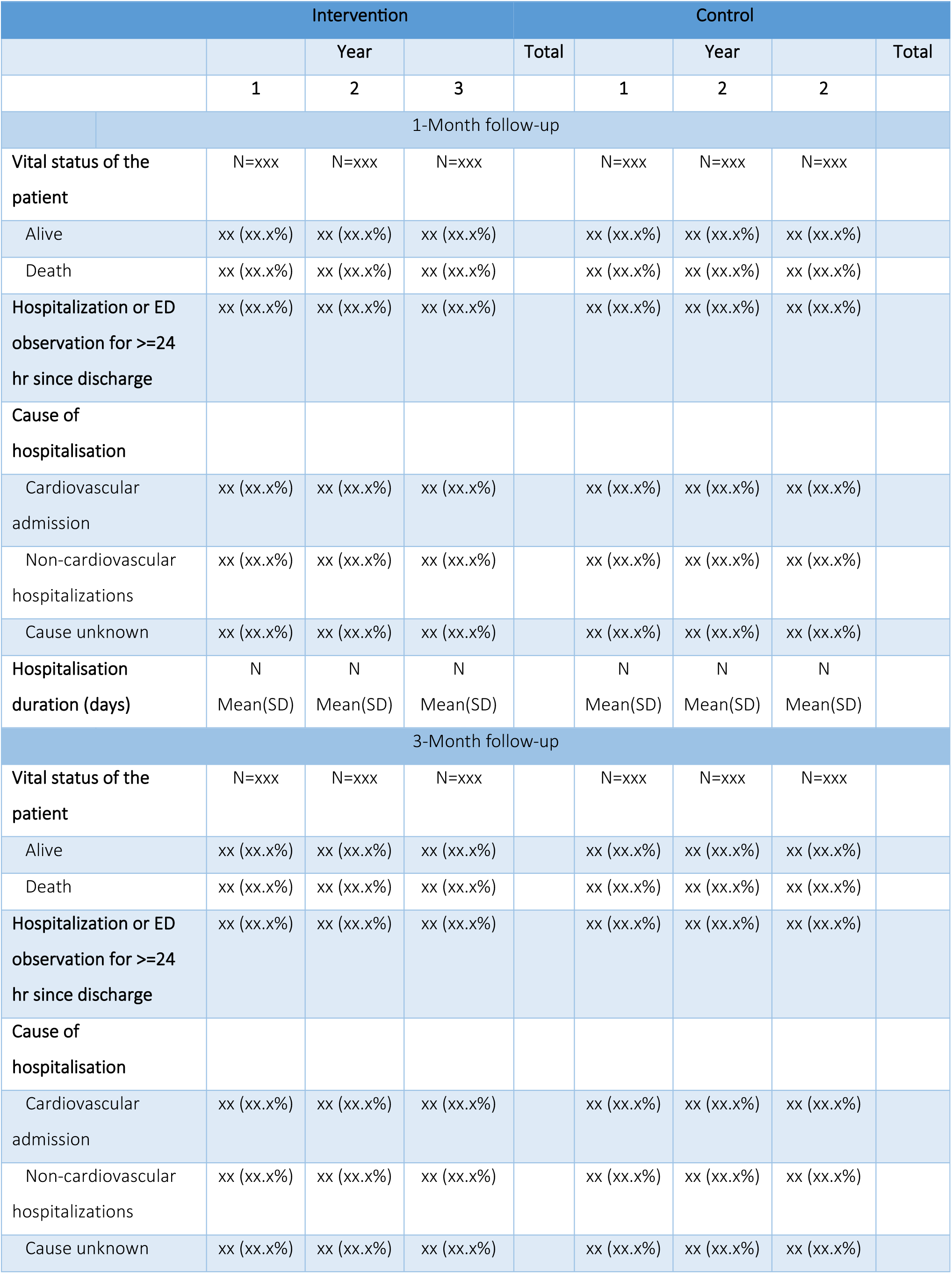
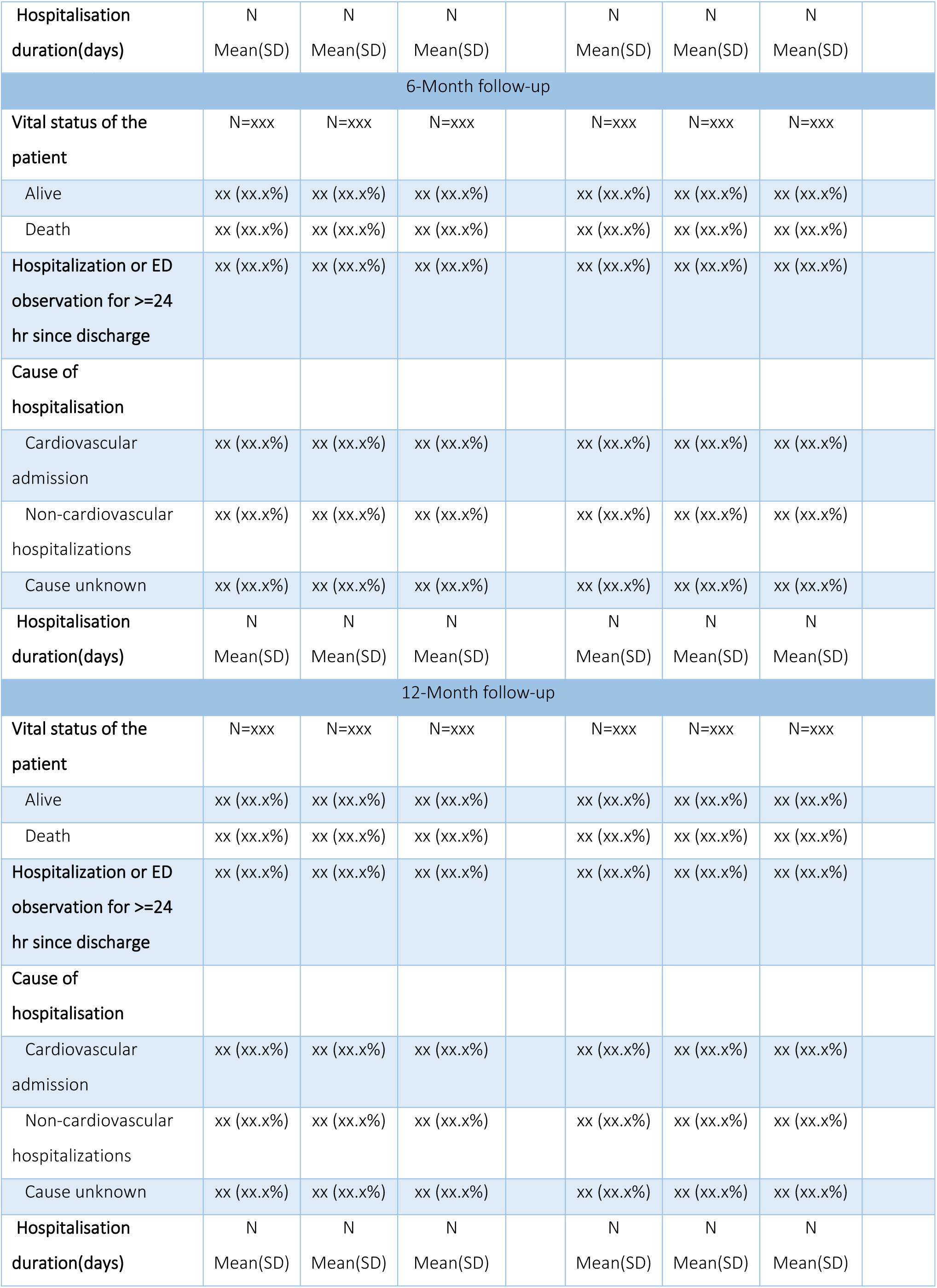
Vital status and hospitalisations by follow-up timepoints and treatment phase.

**Table 15.**
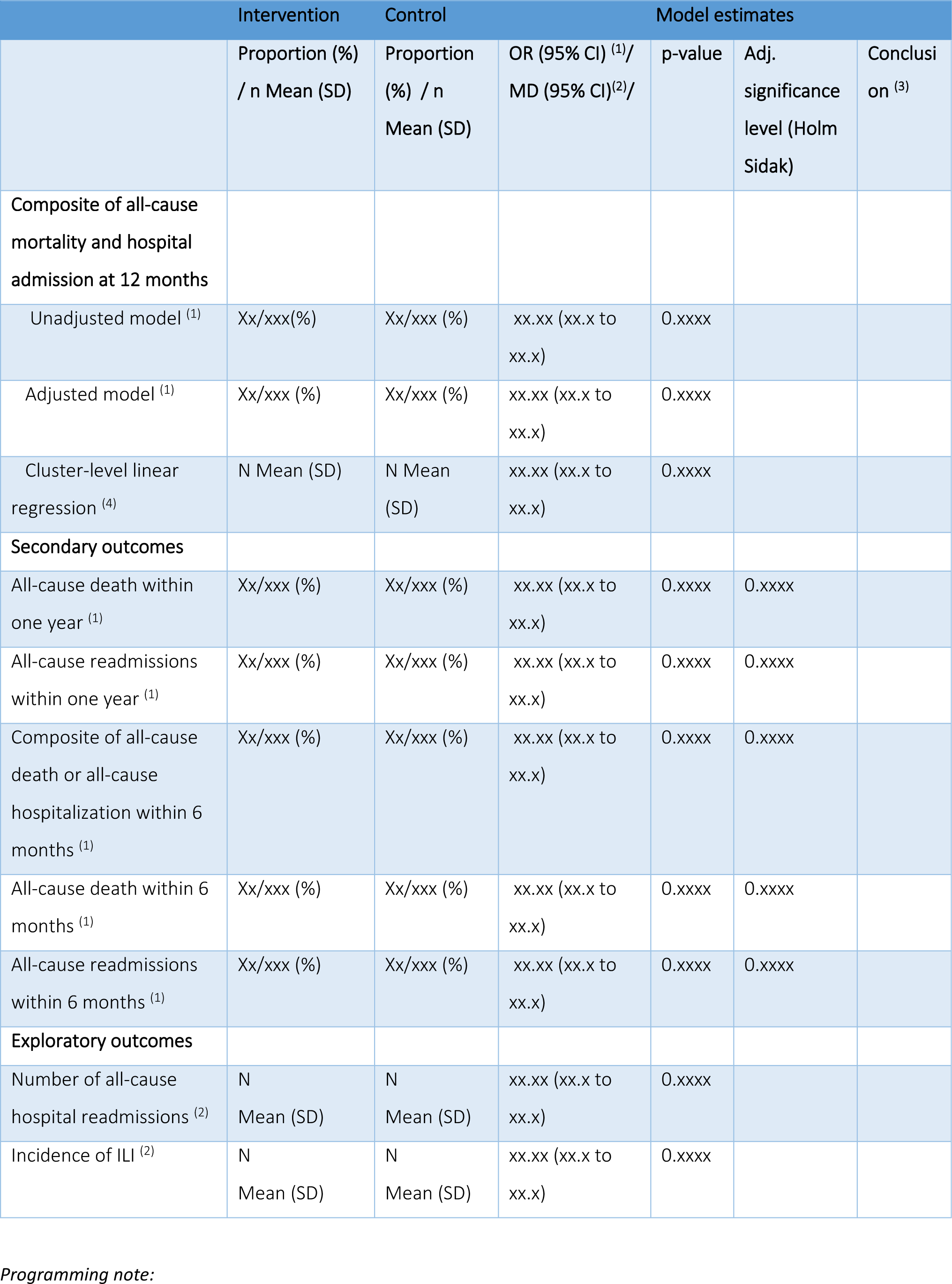
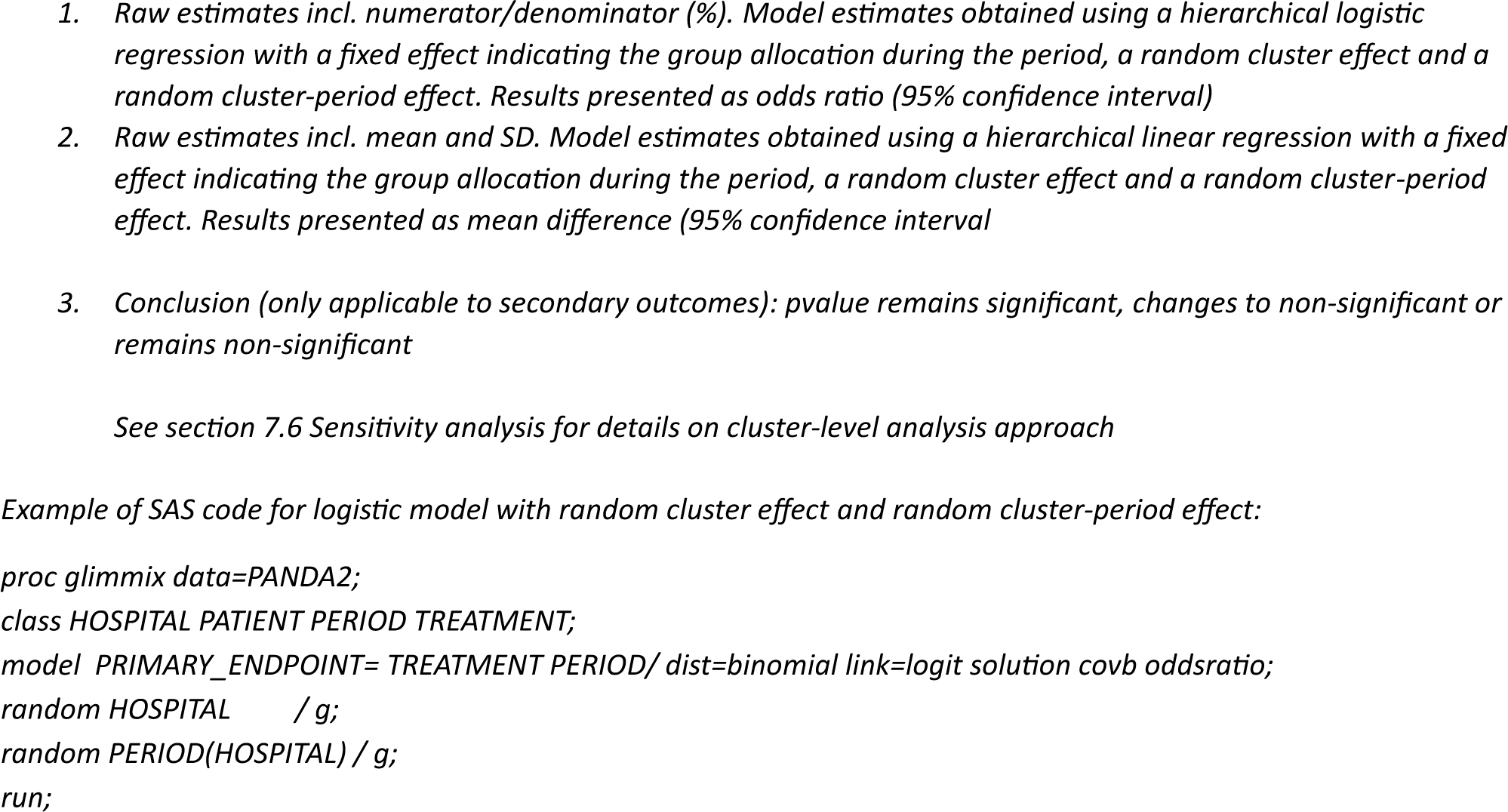
Clinical outcomes – model results.

**Table 16.** Summary of serious adverse events.

|  | Intervention<br>(N =xxx ) | Control<br>(N =xxx ) | p-value<br>(1) |
| --- | --- | --- | --- |
| **Adverse drug reactions after vaccination** | nEVT nPT(xx.x%) | nEVT nPT(xx.x%) | 0.xxx |
| **Serious adverse events (SAE)** | nEVT nPT(xx.x%) | nEVT nPT(xx.x%) | 0.xxx |
| **Serious events** |  |  |  |
| Death or life-threatening | nEVT nPT(xx.x%) | nEVT nPT(xx.x%) |  |
| Causing or prolonging a hospital stay | nEVT nPT(xx.x%) | nEVT nPT(xx.x%) |  |
| Severe disability or incompetence | nEVT nPT(xx.x%) | nEVT nPT(xx.x%) |  |
| Congenital malformations in offspring | nEVT nPT(xx.x%) | nEVT nPT(xx.x%) |  |
| Important clinical events as judged by the investigators | nEVT nPT(xx.x%) | nEVT nPT(xx.x%) |  |
| **SAE severity** |  |  |  |
| Mild (transient, does not interfere with normal activity) | nEVT nPT(xx.x%) | nEVT nPT(xx.x%) |  |
| Moderate (interferes with normal activities) | nEVT nPT(xx.x%) | nEVT nPT(xx.x%) |  |
| Severe (inability to perform daily activities) | nEVT nPT(xx.x%) | nEVT nPT(xx.x%) |  |
| **SAE outcome** |  |  |  |
| Persistent/unresolved | nEVT nPT(xx.x%) | nEVT nPT(xx.x%) |  |
| Resolved but left with sequelae | nEVT nPT(xx.x%) | nEVT nPT(xx.x%) |  |
| Remission | nEVT nPT(xx.x%) | nEVT nPT(xx.x%) |  |
| Fatal | nEVT nPT(xx.x%) | nEVT nPT(xx.x%) |  |
| Unknown | nEVT nPT(xx.x%) | nEVT nPT(xx.x%) |  |
| **Causal relationship to the flu vaccine** |  |  |  |
| Unrelated | nEVT nPT(xx.x%) | nEVT nPT(xx.x%) |  |
| Unlikely to be relevant | nEVT nPT(xx.x%) | nEVT nPT(xx.x%) |  |
| Probably related | nEVT nPT(xx.x%) | nEVT nPT(xx.x%) |  |

1. P-values obtained from cluster-level linear regression as described in section 7.10: Analysis of safety outcomes

**Listing 1. SAE listing**

**Listing 2. AE listing**

**Listing 3. Protocol deviations**

**Figure 2: Forest plot for subgroup analysis of primary outcome**

**Figure 3.**
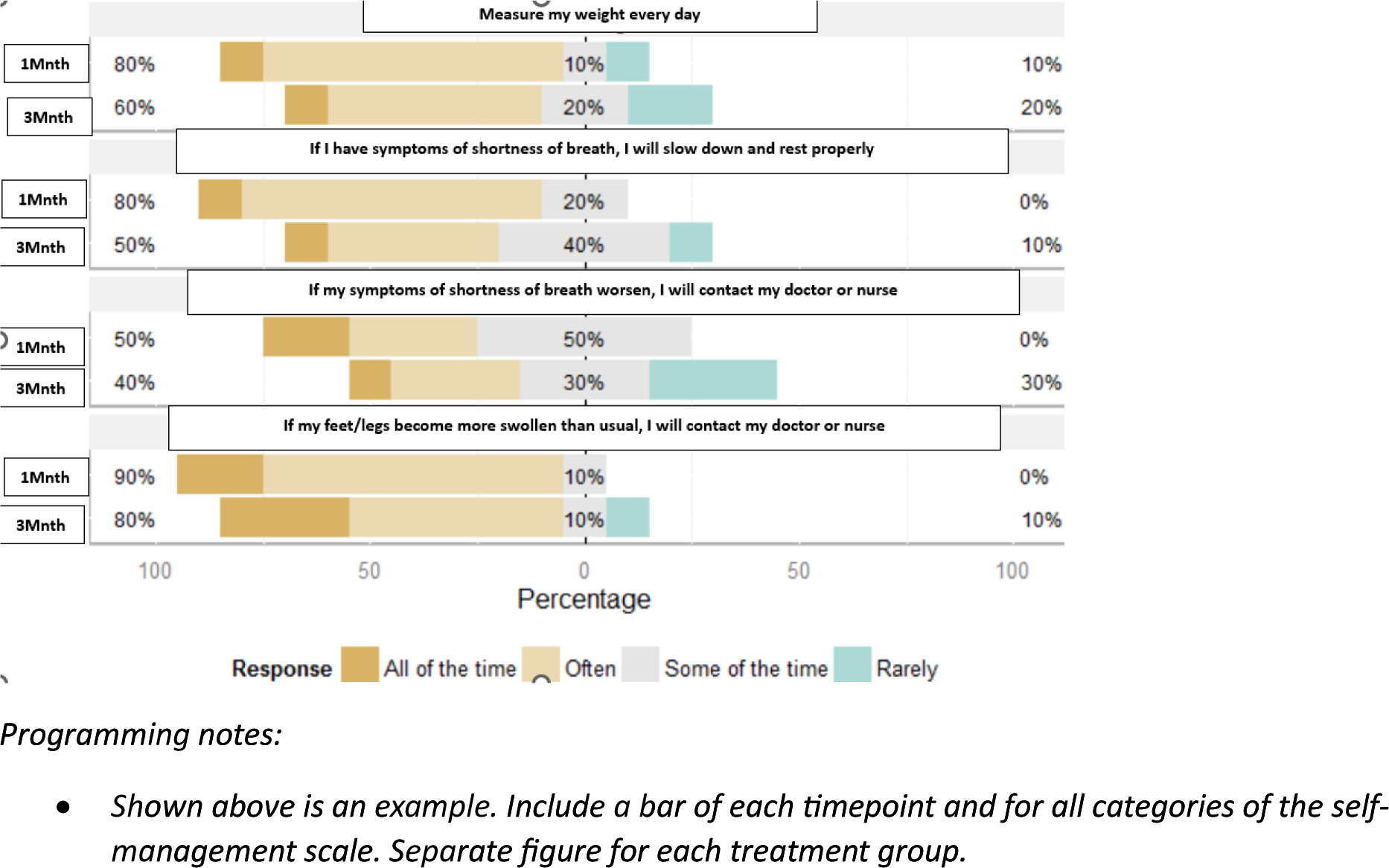
Heart failure self-management scale – descriptive table by timepoints (Month 1, 3, 6, 12)

**Figure 4.**
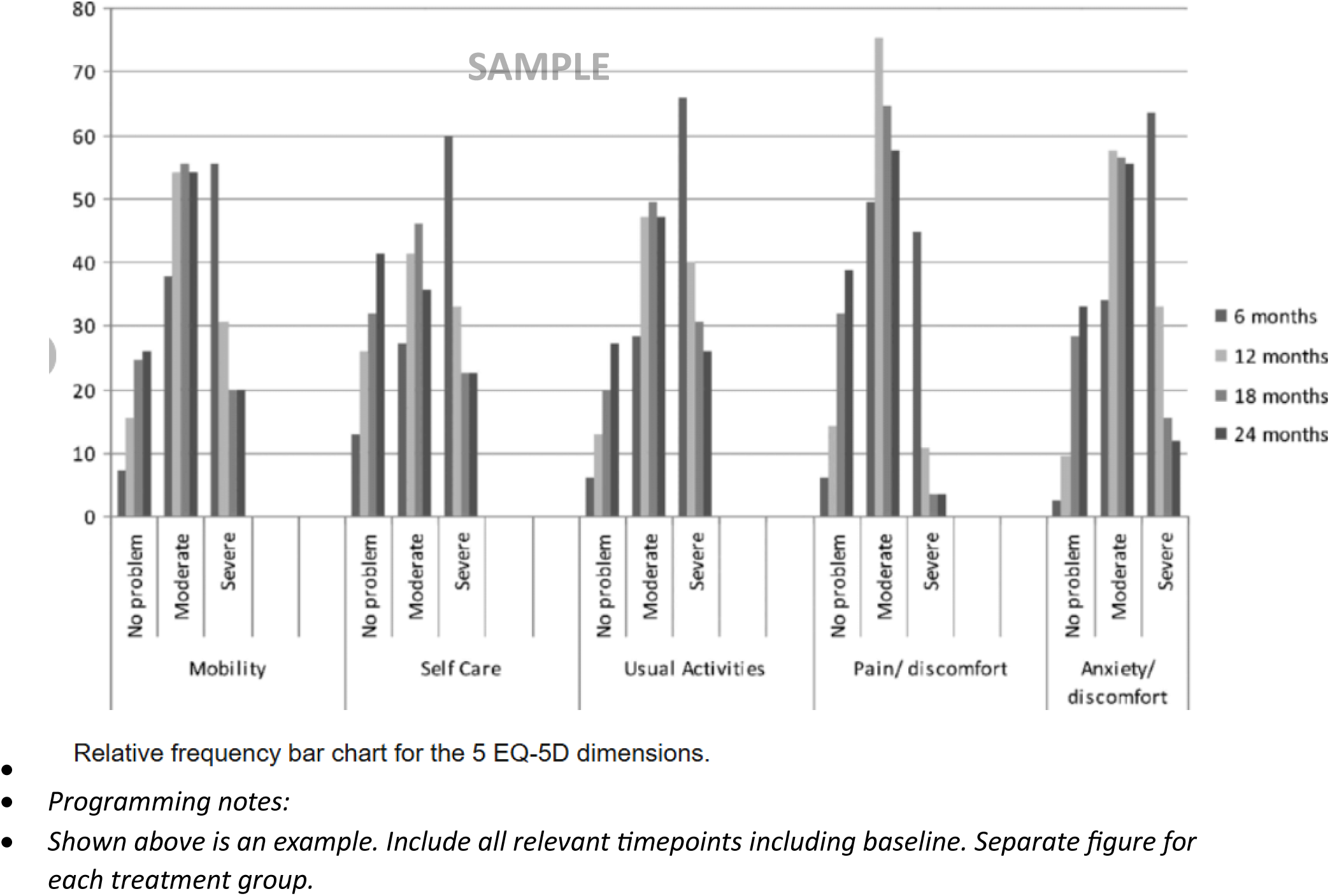
Bar chart of EQ5D-5L across all timepoints (Baseline to 12 months)

**Figure 5. Mean plot of *health utility score (EQ5D)* over time (Baseline to 12 Months) by treatment groups**

*Programming notes:*

*Display raw means on the graph as numbers near each dot and denominators below the x-axis*

